# Cross-cohort insights into the association of handgrip strength transitions and burdens with cardiovascular disease risk

**DOI:** 10.64898/2026.03.05.26347756

**Authors:** Hua Lin, Jike Qi, Yuchen Jiang, Yu Yan, Hao Zhang, Xin Zhang, Jingjing Sun, Min Sun, Wensu Chen, Chen Wang, Tongda Xu, Tianyue Xu, Xingjie Hao, Ting Wang, Chu Zheng, Ke Wang, Ping Zeng

**Author notes:** Co-first authors (Hua Lin, Jike Qi, Yuchen Jiang). **Correspondence to:** Ping Zeng, Department of Biostatistics, Jiangsu Engineering Research Center of Biological Data Mining and Healthcare Transformation, and Jiangsu Key Laboratory of Geriatric Precision Medicine and Aging Intervention, Xuzhou Medical University, Xuzhou, Jiangsu 221004, China.

## Abstract

**Background:** Previous studies established handgrip strength (HGS) as a potential risk factor for cardiovascular diseases (CVD). However, existing studies focused exclusively on baseline HGS and neglected longitudinal changes in HGS during follow-up. Thus, our aim was to investigate the associations of transitions and dynamic burdens of HGS with incident CVD risk.

**Methods:** We analyzed data from the UK Biobank (UKB), the China Health and Retirement Longitudinal Study (CHARLS), the Survey of Health, Ageing and Retirement in Europe (SHARE), and the Korean Longitudinal Study of Ageing (KLOSA). We defined HGS transitions based on HGS at baseline and the first follow-up, and created three indicators to reflect HGS burdens. Cox models were applied to examine the association of HGS transitions and burdens with incident CVD risk. The predictive value of HGS indices was also evaluated.

**Results:** A total of 73,555 participants were retained, and 4,722 (6.4%) incident CVD cases were identified during follow-up. Transition analyses revealed that increased HGS during follow-up was associated with reduced CVD risk, whereas decreased HGS was associated with an elevated risk. Per standard deviation decrement in HGS slope, cumulative HGS and relative cumulative HGS led to a 19.8% (95%CI 1.5~41.5%), 44.0% (95%CI 10.8~87.2%) or 26.7% (95%CI 9.4~46.8%) elevated risk of CVD, respectively. These associations were independent of and more pronounced than HGS, with stronger effects observed in East Asian cohorts (CHARLS and KLOSA) compared to European cohorts (UKB and SHARE). Incorporating dynamic HGS metrics enhanced the predictive accuracy, with HGS burdens providing greater gains than HGS. For the optimal models, all HGS indices resulted in an increase of AUC up to 7.6% in Europeans and 5.9% in East Asians.

**Conclusions:** HGS burdens outperformed in predicting cardiovascular health compared to single cross-sectional HGS itself, suggesting the clinical utility of longitudinal HGS monitoring in clinical and public health strategies for CVD prevention.

## Introduction

Cardiovascular disease (CVD) remains the leading cause of death and a significant contributor to the global burden of disease (Roth, et al., 2020), making early prevention essential for improving population health and reducing disease burden. Handgrip strength (HGS), a simple and non-invasive measurement of muscle strength, has garnered increasing attention as a potential predictive indicator of aging and disease risk because of its ease of operation, low cost, and high reproducibility (Duchowny, et al., 2022; Jiang, et al., 2022; Kuo, et al., 2023; Yu, et al., 2025; Zhang, et al., 2024). Substantial evidence has shown associations between HGS decline and many adverse health outcomes, including type 2 diabetes (Marcotte-Chénard, et al., 2023), chronic kidney disease (Zhang, et al., 2025), and cancer (López-Bueno, et al., 2022). Particularly, lower HGS is associated with greater incidence of specific CVDs, such as stroke (Gao, et al., 2022; Leong, et al., 2015), arrhythmia (Andersen, et al., 2015), and coronary heart disease (Ding, et al., 2020), and has become an early signal of the pathophysiologic processes underlying CVD.

Although the exact mechanism is not fully understood, the association between HGS and CVD may involve the muscle-cardiovascular crosstalk. Reduced HGS likely results from lasting chronic low-grade inflammation (Kang, et al., 2021), metabolic disturbances (Li, et al., 2018; Wu, et al., 2025), mitochondrial dysfunction (Kothari, et al., 2024), and aging (Kemala Sari, et al., 2025; Vaishya, et al., 2024; Vennu, 2023), which concurrently lead to the development of CVD. The relationship between declining HGS and rising CVD risk is further amplified by inflammatory mediators (Cesari, et al., 2004). For instance, chronic low-grade inflammation and hormonal dysregulation create a vicious cycle that simultaneously compromises skeletal muscle and cardiovascular health (Xie, et al., 2023).

However, currently HGS has not yet been interpreted or applied in a uniform manner. Some studies employed the maximal or mean HGS of individuals′ dominant hand, while others calculated HGS by summing the maximal HGS of both hands (Cheung, et al., 2012; Mainous, et al., 2015; van der Kooi, et al., 2015), which may be confounded by an individual′s weight, height, and other physical fitness indicators. To account for the influences of these factors on HGS (Hardy, et al., 2013), the Foundation of the United States National Institutes of Health has proposed a new HGS metric to correct for body size by dividing muscle strength by weight (Studenski, et al., 2014). This adjusted HGS, also called relative HGS to be distinguished from the absolute HGS, is comparable across different measurement approaches and enhances its translational value as a prognostic tool (Lawman, et al., 2016). Several studies have demonstrated that the adjusted HGS is better predictive for the risk of cardio-metabolic morbidity (Lee, et al., 2016; Li, et al., 2018; Yi, et al., 2018) and mortality (Dong, et al., 2016) compared to its unadjusted counterpart.

Although a growing body of research has explored the association between this type of HGS and CVD risk from various perspectives (Chi and Lee, 2024; Kim, et al., 2021; Xie, et al., 2023), there still exists room for further investigation. First, since HGS changes over time and previous studies relied primarily on a single cross-sectional HGS measurement at baseline to predict CVD outcomes (Laukkanen, et al., 2020; Leong, et al., 2015; Liu, et al., 2021), large-scale long-term longitudinal cohorts with repeated visits are lacking to capture the actual dynamic trajectory of HGS. Therefore, the understanding of the association between HGS change patterns and cardiovascular risk remains at a static level; whereas cumulative and dynamic changes in HGS often better reflect the long-term state of muscle metabolic function (Wen, et al., 2022), and persistent exposure to muscle metabolic disorders (such as insulin resistance) is an important driver of CVD (Sun, et al., 2025). Second, most findings originated from a single population cohort (Prasitsiriphon and Pothisiri, 2018; Qiu, et al., 2023; Yang, et al., 2024), the lack of sufficient cross-population comparisons and validations in different cohorts limits the generalizability of current results. Existing studies have implied that the impact of HGS on CVD differs between European and East Asian populations; for instance, a per-unit decrease in HGS was linked to a 7% greater CVD risk among Europeans (López-Bueno, et al., 2022), whereas it was about 17% in Asians (Leong, et al., 2015). However, there are few multi-cohort studies to date to verify the statistical significance of the racial difference. Finally, current studies often employed only a single dynamic indicator (e.g., slope) to assess the association of dynamic changes in HGS with CVD, failing to systematically integrate and compare multiple dynamic indicators that reflect different patterns of change (Bae, et al., 2021; Qiu, et al., 2023; Zammit, et al., 2021). Overall, this requires a more comprehensive and in-depth analysis of the complex relationship between dynamic changes in HGS and CVD risk.

We here examined the connection between HGS and CVD risk within four regionally representative prospective cohorts, including the UK Biobank (UKB) (Sudlow, et al., 2015), the China Health and Retirement Longitudinal Study (CHARLS) (Zhao, et al., 2014), the Survey of Health, Ageing and Retirement in Europe (SHARE) (Andersen-Ranberg, et al., 2009), and the Korean Longitudinal Study of Ageing (KLOSA) (Min, et al., 2012). Each of these cohorts has its own unique characteristics; their integration not only increases sample sizes but also provides an opportunity for cross-population verification of the HGS-CVD relationship. We employed the adjusted HGS to assess their temporal associations with CVD events across the four cohorts, and defined three HGS burden indicators (HGS slope, cumulative HGS, and relative cumulative HGS) to evaluate how the dynamic change in HGS affected the risk of CVD. These three indicators correspond to the dynamic change rate, total amount and relative change, respectively, which reflect the different dimensions of exposure in a complementary way and have been proven effective in other diseases (He, et al., 2024; Zhang, et al., 2025). We finally assessed whether these HGS metrics would offer additional predictive value when incorporating them into SCORE2 (SCORE2 working group and ESC Cardiovascular risk collaboration, 2021), one of the most widely used cardiovascular risk assessment algorithms, which currently lacks sufficient capture of risk factors related to muscle function.

## Methods and Materials

### Data source and study design

We analyzed data from the UKB (Sudlow, et al., 2015), CHARLS (Zhao, et al., 2014), SHARE (Andersen-Ranberg, et al., 2009) and KLOSA (Min, et al., 2012) cohorts, each of which contained waves with HGS measurements at baseline (the first time at which HGS was measured) and during follow-up (Figure 1). To enhance homogeneity and analytical robustness, our study was restricted to participants of White European and East Asian ancestry. Baseline assessments (*t*_0_) included UKB wave 1 (2006-2010), CHARLS wave 1 (2011), SHARE wave 6 (2015), and KLOSA wave 3 (2010). The first follow-up (*t*_1_) comprised UKB wave 2 (2012-2014), CHARLS wave 2 (2013), SHARE wave 7 (2017), and KLOSA wave 4 (2012), with an average interval time of approximately two years in every cohort. The last follow-up included UKB wave 3 (2021), CHARLS wave 4 (2018), SHARE wave 9 (2021), and KLOSA wave 7 (2018).

**Figure 1.**
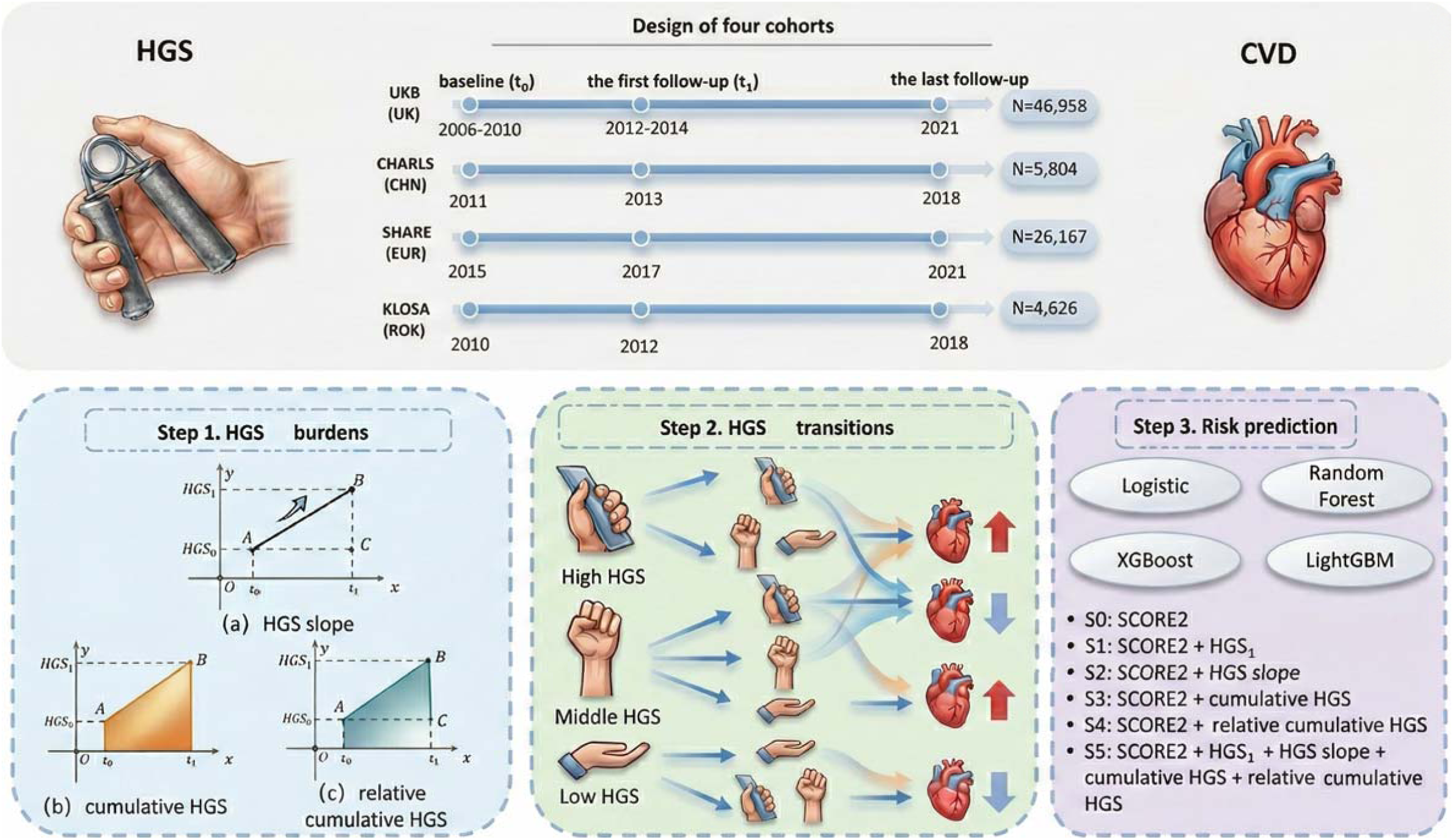
Study design overview for investigating the relationship between HGS and CVD. CVD: cardiovascular disease. HGS: handgrip strength; HGS_0_ and HGS_1_ refer to HGS for each participant measured at baseline and the first follow-up, respectively.

We excluded individuals who met the following characteristics: (i) missing left- or right-hand measurements of HGS; (ii) previous diagnosis of CVD at baseline; (iii) loss to follow-up or withdrawal of informed consent during the study period. We ultimately included 73,555 individuals, of whom 46,958 participants from UKB, 5,804 from CHARLS, 26,167 from SHARE, and 4,626 from KLOSA (Figure S1).

### Definition of CVD and handgrip strength

To obtain sufficient disease cases to enhance statistical power, incident CVDs were defined as composite events of coronary heart disease, heart failure, atrial fibrillation, or stroke (Tikkanen, et al., 2018). In UKB, diagnoses were obtained from hospital records and death registers (including England, Scotland, and Wales), and recorded using ICD-9 and ICD-10 (Table S1); whereas in the other three databases, CVD cases were defined as a history of stroke and heart problems. The endpoint was the date of diagnosis of CVD events, death, loss to follow-up, or visit deadline, whichever occurred first.

HGS was measured consistently across the four cohorts using a Jamar 100105 hydraulic hand dynamometer. Participants were seated upright with elbows at their sides and forearms bent at 90° on armrests. Isometric HGS was measured by a single 3-second maximal squeeze for each hand (Choquette, et al., 2010; Ho, et al., 2019; Lawman, et al., 2016). Given the established association between muscle strength and body mass (Peterson, et al., 2016), we defined HGS as the mean of left- and right-hand measurements adjusted for body weight; this adjusted HGS largely balances the differences in HGS among various races and populations (Bhat, et al., 2021; Dodds, et al., 2016).

### Selection of covariates

We selected the following covariates based on their availability from the four cohorts: age at recruitment, sex, socioeconomic factors (e.g., educational level and income), behavioral factors (e.g., alcohol consumption status, smoking status and physical activity status), health status (e.g., diabetes and hypertension), and body mass index (BMI) (D’Agostino, et al., 2008; Magnussen, et al., 2023; Samarasekera, et al., 2023). Multiple imputation was performed via the multivariate imputation by chained equations (MICE) method to impute missing values for each covariate (the maximal missing rate was 16% for physical activity in KLOSA) (van Buuren and Groothuis-Oudshoorn, 2011).

### Definition of HGS transitions and burdens

Based on the categories of low, moderate, and high HGS defined by tertiles at baseline and the first measurement, HGS transitions were divided into seven categories: low HGS → low HGS, low HGS → increased HGS (including moderate HGS and high HGS for improving sample size in this category), moderate HGS → moderate HGS, moderate HGS → high HGS, moderate HGS → low HGS, high HGS → high HGS, and high HGS → decreased HGS (including moderate HGS and low HGS for improving sample size in this category).

To quantify the HGS burden, we constructed three continuous indicators (Cui, et al., 2022; He, et al., 2024; Xiang, et al., 2024; Zhang, et al., 2025):

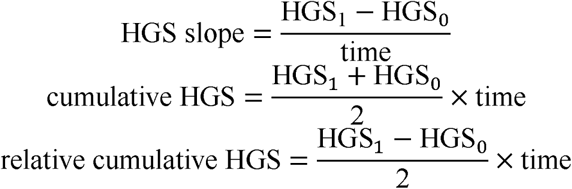

where HGS_0_ and HGS_1_ stand for HGS measured at baseline and at the first follow-up, time (years) is the interval between the baseline and the first follow-up assessments (i.e., *t*_1_-*t*_0_ in Figure 1). HGS slope reflects the rate of HGS change; cumulative HGS indicates the area under the HGS vs. time trajectory; relative cumulative HGS shows cumulative HGS that has increased or decreased relative to baseline HGS. These three burden indicators represent distinct dimensions of dynamic changes in exposure and have been successfully employed in previous studies (He, et al., 2024).

### Statistical analyses

#### Association of HGS transitions with incident CVD risk

We first assessed the impact of dynamic changes in HGS on CVD risk by establishing three reference groups for comparative analysis. Specifically, among participants with initially high HGS, those who consistently maintained high HGS (denoted as high → high) were designated as the reference group; comparisons were made against those whose HGS declined to a lower category. In the moderate HGS group, individuals with stable moderate HGS (moderate → moderate) served as the reference, and we examined transitions either to low (moderate → low) or to high (moderate → high) HGS. Similarly, among those with low baseline HGS, participants who remained in the low HGS category (low → low) were used as the reference, and those showing improvement to higher HGS levels were analyzed.

Covariate-adjusted Cox proportional hazards (PH) models were employed to evaluate the correlation between HGS and incident CVD risk in each transition. Results were expressed as hazard ratio (HR) along with its 95% confidence interval (95% CI) for standardized HGS and its burden indicators. No evidence of violation of the PH assumption was observed according to Schoenfeld′s residuals (*P*>0.05). Due to limited samples in some transition subgroups, we merged the four cohorts for stability and robustness, but also differentiated between East Asian (CHARLS and KLOSA) and European (UKB and SHARE) analytical cohorts.

#### Association of HGS burden with incident CVD risk

Next, we used covariate-adjusted Cox model to evaluate the link between each HGS burden and incident CVD risk separately in every cohort. Here, HGS itself was also adjusted to examine the independent role of HGS burden (Sun, et al., 2024). We then pooled individual effect sizes via a random-effect or fixed-effect meta-analysis, depending on the presence of effect heterogeneity across cohorts according to Cochran′s Q statistic.

#### Sensitivity and subgroup analyses

To validate the robustness of our results, we conducted multiple sensitivity analyses: (i) to minimize the reverse causality, we excluded participants who developed CVD during the first two years after the first follow-up; (ii) to evaluate the impact of drug treatments, we removed participants who underwent antihypertensive, statin, or insulin treatments at baseline and the first follow-up; (iii) to directly address competing risks from all-cause deaths, we conducted the Fine-Gray model (Fine and Gray, 1999), reducing potential bias in standard Cox model estimates. Considering that age is an important risk factor for CVD (Zhao, et al., 2024) and HGS levels correlate with sex in the risk of disease development (Li, et al., 2023), we carried out subgroup analyses stratified by age groups (<65 or ≥65 years) and sex (female or male).

#### Evaluation of predictive capacity for HGS

To evaluate the predictive value of HGS combined with SCORE2 (Table S2), six scenarios with different predictors were considered: S0 (base model): SCORE2; S1: SCORE2 + HGS; S2: SCORE2 + HGS slope; S3: SCORE2 + cumulative HGS; S4: SCORE2 + relative cumulative HGS; S5: SCORE2 + HGS + HGS slope + cumulative HGS + relative cumulative HGS. Given the evident racial heterogeneity in effects of HGS, we carried out the prediction analysis separately in European and East Asian cohorts, excluding KLOSA due to missing blood sample data required for SCORE2 calculation.

In each scenario, we implemented four machine learning models (logistic regression, randomForest, XGBoost, and LightGBM) via 10 replications of Monte Carlo cross validation for internal validation (80% training, 20% test) (Zeng and Zhou, 2017). External validation was also performed on an independent cohort to assess generalizability. For example, models trained in UKB and SHARE were validated in CHARLS, and *vice versa*. The AUC and its 95%CI were used to evaluate model prediction performance.

To assess the improvement in risk classification, we calculated the net reclassification improvement (NRI) and the integrated discrimination improvement (IDI), using a CVD risk threshold of 7.5% to define high-risk individuals (Grundy, et al., 2019). These metrics allowed us to quantify the proportion of cases and non-cases correctly reclassified by the enhanced models, as well as the overall improvement in the model’s discriminatory power.

#### Statistical software

All analyses were conducted within the R (version 4.3.0) statistical computing environment. Missing values were imputed via the mice package (van Buuren and Groothuis-Oudshoorn, 2011). Cox models were implemented using the survival package (version 3.8.3). For predictive modeling, we relied on the randomForest (version 4.7.1.2) (Liaw and Wiener, 2007), xgboost (version 1.7.10.1) (Chen and Guestrin, 2016), and lightgbm (version 4.6.0) (Ke, et al., 2017) packages. A two-tailed *P*-value < 0.05 was considered statistically significant.

## Results

### Basic characteristics of the four cohort studies

We ascertained 73,555 participants (Table 1), including 46,958 from UKB (female: 56.0%, mean age: 63.0±7.8 years), 5,804 from CHARLS (female: 53.7%, mean age: 60.8±9.4 years), 26,167 from SHARE (female: 56.6%, mean age: 67.3±9.6 years), and 4,626 from KLOSA (female: 55.3%, mean age: 64.4±9.0 years). The median follow-up time for UKB, CHARLS, SHARE, and KLOSA was 8.8 years (IQR: 3.6~12.1 years), 5.0 years (IQR: 1.9~7.1 years), 4.5 years (IQR: 1.6~5.8 years), and 6.7 years (IQR: 2.4~7.5 years), respectively. During the follow-up period, a total of 4,722 (6.4%) incident CVD cases were identified.

**Table 1.**
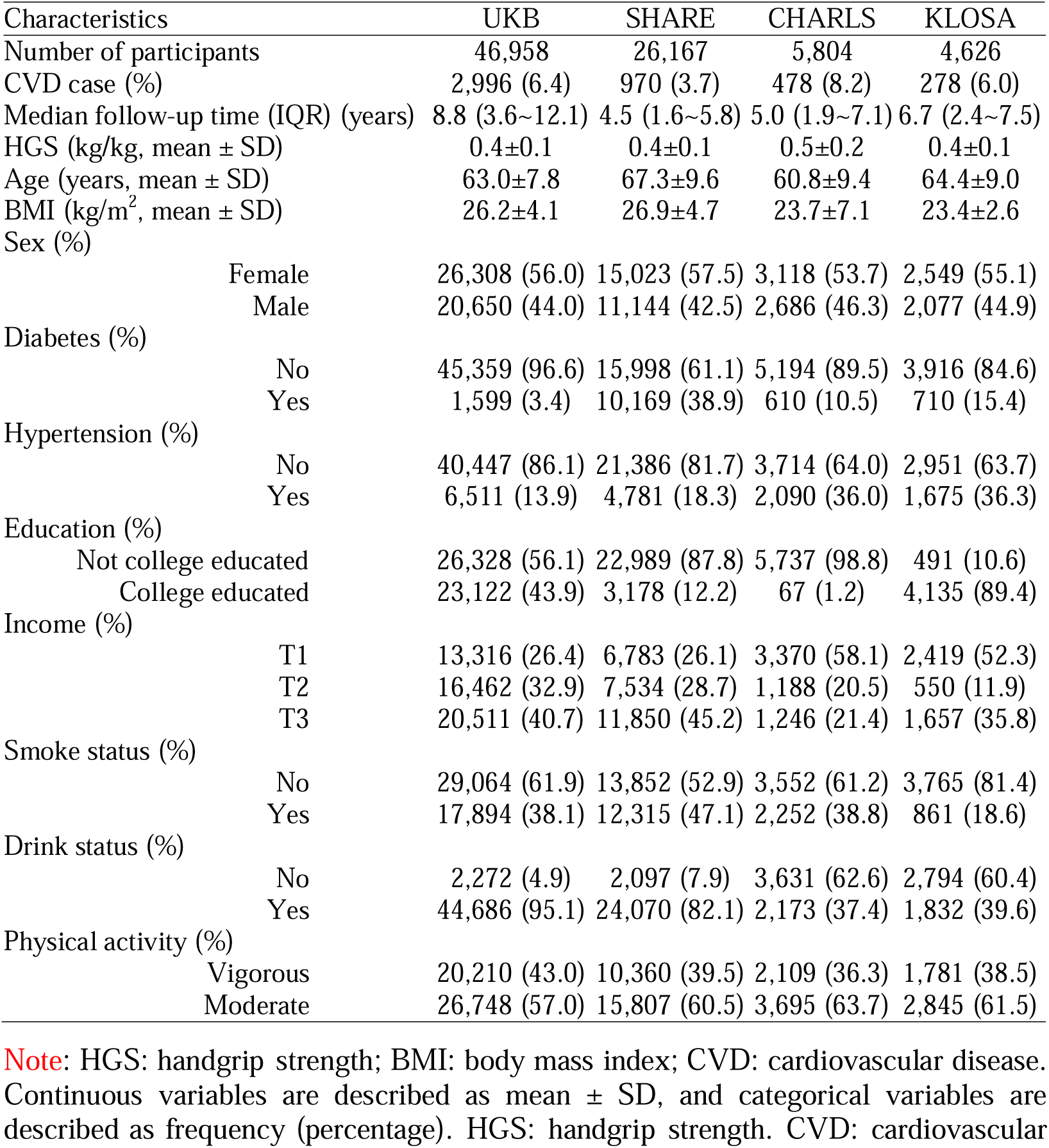
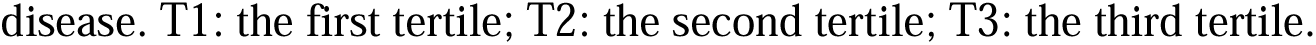
Characteristics of participants from the four cohorts.

| Characteristics | UKB | SHARE | CHARLS | KLOSA |
| --- | --- | --- | --- | --- |
| Number of participants | 46,958 | 26,167 | 5,804 | 4,626 |
| CVD case (%) | 2,996 (6.4) | 970 (3.7) | 478 (8.2) | 278 (6.0) |
| Median follow-up time (IQR) (years) | 8.8 (3.6~12.1) | 4.5 (1.6~5.8) | 5.0 (1.9~7.1) | 6.7 (2.4~7.5) |
| HGS (kg/kg, mean ± SD) | 0.4±0.1 | 0.4±0.1 | 0.5±0.2 | 0.4±0.1 |
| Age (years, mean ± SD) | 63.0±7.8 | 67.3±9.6 | 60.8±9.4 | 64.4±9.0 |
| BMI (kg/m<sup>2</sup>, mean ± SD) | 26.2±4.1 | 26.9±4.7 | 23.7±7.1 | 23.4±2.6 |
| Sex (%) |  |  |  |  |
| Female | 26,308 (56.0) | 15,023 (57.5) | 3,118 (53.7) | 2,549 (55.1) |
| Male | 20,650 (44.0) | 11,144 (42.5) | 2,686 (46.3) | 2,077 (44.9) |
| Diabetes (%) |  |  |  |  |
| No | 45,359 (96.6) | 15,998 (61.1) | 5,194 (89.5) | 3,916 (84.6) |
| Yes | 1,599 (3.4) | 10,169 (38.9) | 610 (10.5) | 710 (15.4) |
| Hypertension (%) |  |  |  |  |
| No | 40,447 (86.1) | 21,386 (81.7) | 3,714 (64.0) | 2,951 (63.7) |
| Yes | 6,511 (13.9) | 4,781 (18.3) | 2,090 (36.0) | 1,675 (36.3) |
| Education (%) |  |  |  |  |
| Not college educated | 26,328 (56.1) | 22,989 (87.8) | 5,737 (98.8) | 491 (10.6) |
| College educated | 23,122 (43.9) | 3,178 (12.2) | 67 (1.2) | 4,135 (89.4) |
| Income (%) |  |  |  |  |
| T1 | 13,316 (26.4) | 6,783 (26.1) | 3,370 (58.1) | 2,419 (52.3) |
| T2 | 16,462 (32.9) | 7,534 (28.7) | 1,188 (20.5) | 550 (11.9) |
| T3 | 20,511 (40.7) | 11,850 (45.2) | 1,246 (21.4) | 1,657 (35.8) |
| Smoke status (%) |  |  |  |  |
| No | 29,064 (61.9) | 13,852 (52.9) | 3,552 (61.2) | 3,765 (81.4) |
| Yes | 17,894 (38.1) | 12,315 (47.1) | 2,252 (38.8) | 861 (18.6) |
| Drink status (%) |  |  |  |  |
| No | 2,272 (4.9) | 2,097 (7.9) | 3,631 (62.6) | 2,794 (60.4) |
| Yes | 44,686 (95.1) | 24,070 (82.1) | 2,173 (37.4) | 1,832 (39.6) |
| Physical activity (%) |  |  |  |  |
| Vigorous | 20,210 (43.0) | 10,360 (39.5) | 2,109 (36.3) | 1,781 (38.5) |
| Moderate | 26,748 (57.0) | 15,807 (60.5) | 3,695 (63.7) | 2,845 (61.5) |

Note: HGS: handgrip strength; BMI: body mass index; CVD: cardiovascular disease. Continuous variables are described as mean ± SD, and categorical variables are described as frequency (percentage). HGS: handgrip strength. CVD: cardiovascular
disease. T1: the first tertile; T2: the second tertile; T3: the third tertile.

### Association of HGS transitions with incident CVD risk

Participants who transitioned from low to increased HGS showed a 28.3% (95%CI 13.9~38.1%) lower CVD risk compared to those maintaining stable low HGS (Figure 2). Compared to participants remaining stable moderate HGS, those shifting from moderate to increased HGS demonstrated an 8.4% (95%CI 2.8~14.1%) lower risk. Conversely, participants who shifted from moderate to decreased HGS encountered a 19.3% (95%CI 2.2~38.7%) higher risk of CVD, and those transitioning from high to decreased HGS had a 31.2% (95%CI 16.5~49.6%) elevated risk compared to those who maintained a stable high HGS. Overall, increased HGS during follow-up was associated with reduced CVD risk, whereas decreased HGS was linked to elevated risk.

**Figure 2.**
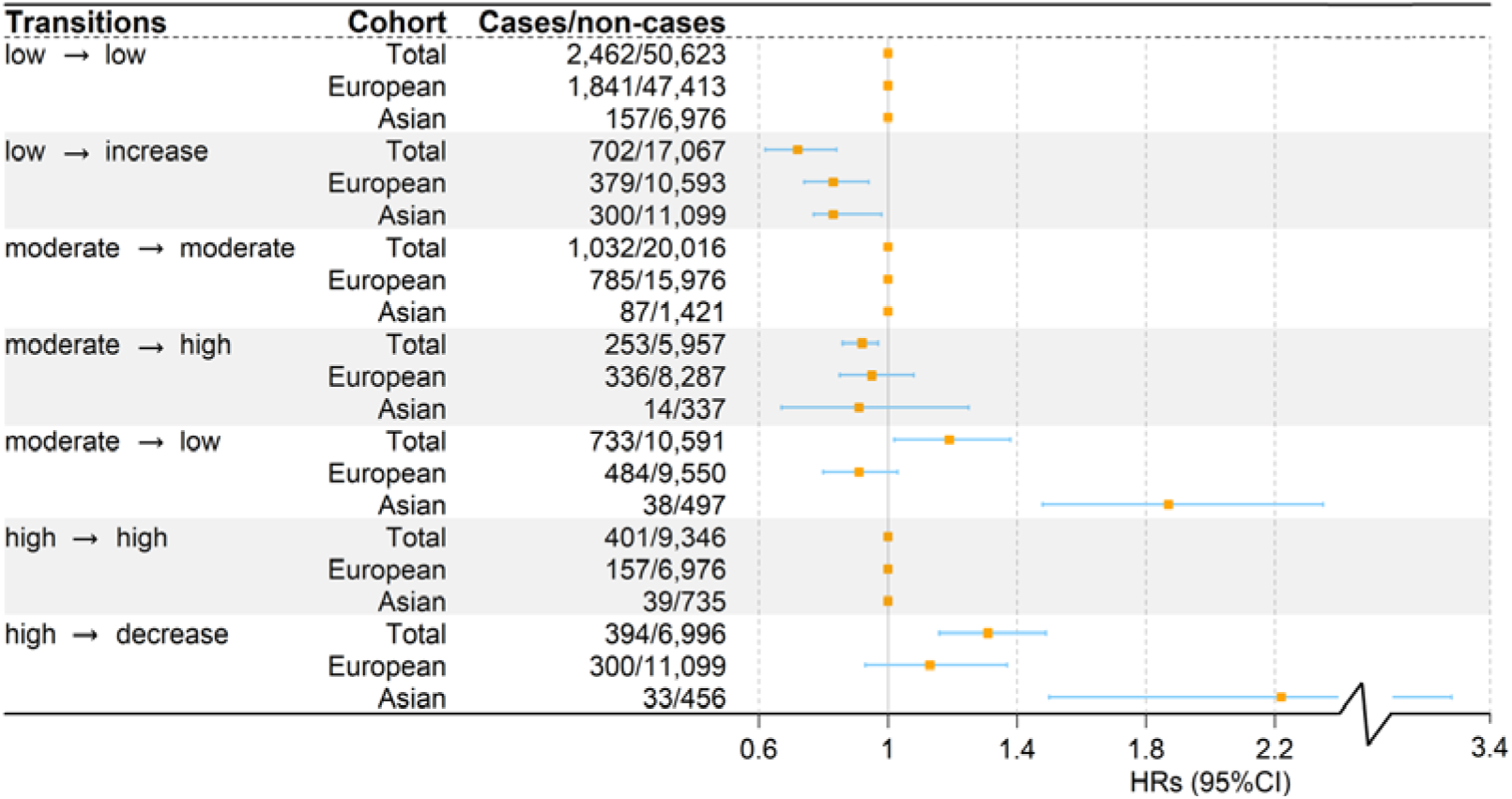
Forest plots of association of HGS transitions with the incident CVD risk. HGS: handgrip strength. CVD: cardiovascular disease.

To evaluate potential ethnic differences, transition analysis was conducted separately for East Asian and European cohorts (Figure 2). Notably, the cardiovascular risk increased by 87.6% (95%CI 48.1~135.5%) in East Asian cohorts when participants transitioned from moderate to low HGS, whereas this association was not significant in European cohorts (*P*=0.156). Similarly, when transitioning from high to decreased HGS, the risk increased by 122.2% (95%CI 50.7~228.1%) in East Asian cohorts, compared to an elevated but non-significant risk of 13.3% (95%CI −7.1~37.8%) in European cohorts.

### Association of HGS burdens with incident CVD risk

After explaining the influence of HGS, we found that HGS burdens played an independent role in the development of CVD (Figure 3). Overall, across the four cohorts, each SD decrement in HGS slope was associated with a 19.8% (95%CI 1.5~41.5%) higher risk of CVD, while each SD decrement in cumulative HGS and relative cumulative HGS led to a 44.0% (95%CI 10.8~87.2%) and 26.7% (95%CI 9.4~46.8%) increased CVD risk, respectively.

**Figure 3.**
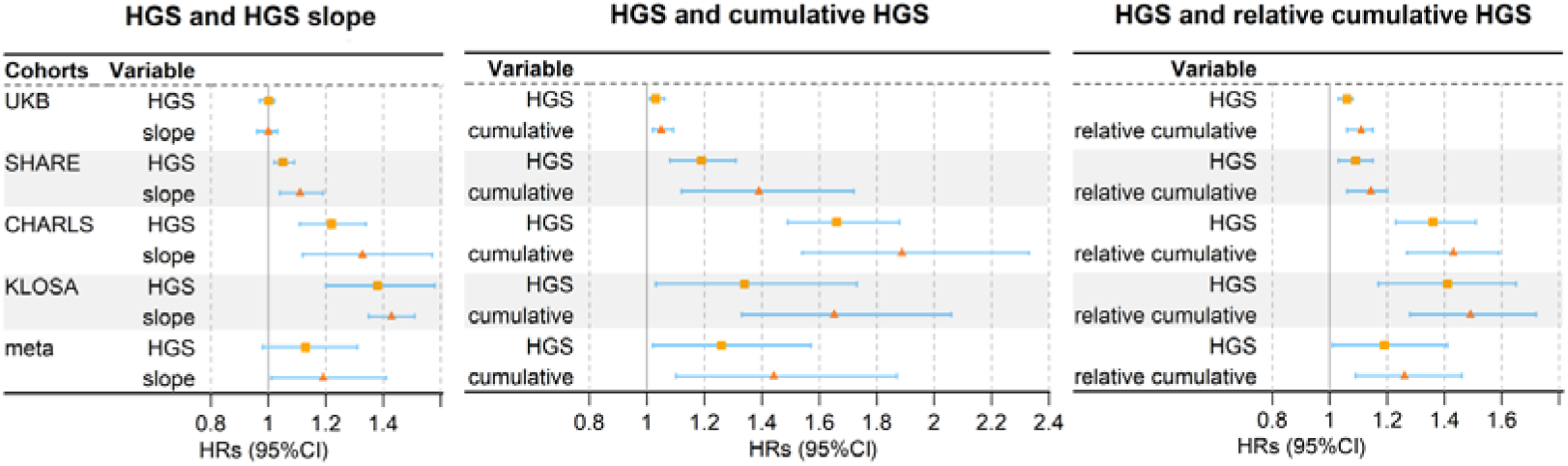
Forest plot illustrates the relationship between HGS and HGS burdens with incident CVD risk. HGS: handgrip strength. CVD: cardiovascular disease.

These associations demonstrated several important patterns. First, the impact of each HGS burden displayed significant heterogeneity across cohorts (*P*<0.0001), with the influence being more evident in East Asian cohorts (KLOSA and CHARLS) than in European cohorts (UKB and SHARE). For instance, for HGS slope, we detected more pronounced risk increase of 43.1% (95%CI 35.7~51.4%) in KLOSA and 33.2% (95%CI 12.7~57.4%) in CHARLS, while observing a lower elevation of 11.2% (95%CI 4.9~19.5%) in SHARE and a non-significant change of −1.2% (95%CI −4.1~3.5%) in UKB. Similar patterns were identified for cumulative and relative cumulative HGS. Second, compared to HGS slope and relative cumulative HGS, each SD decrement of cumulative HGS generally resulted in a higher risk of CVD across all cohorts.

Importantly, for each SD reduction, HGS burdens typically demonstrated a stronger risk effect on CVD compared to HGS (HR=1.19 [95%CI 1.01~1.41] vs. 1.13 [95%CI 0.98~1.31] for HGS slope, 1.44 [95%CI 1.10~1.87] vs. 1.26 [95%CI 1.02~1.57] for cumulative HGS, 1.26 [95%CI 1.09~1.46] vs. 1.19 [95%CI 1.01~1.41] for relative cumulative HGS). This pattern was consistently observed in each cohort (Figure 3). In a further analysis conducted only in UKB, the analogous pattern was also observed for individual CVDs; that is, for each SD decrease, HGS burdens exhibited stronger risk associations than HGS (Figure S2). In addition, the impacts of HGS burdens were especially pronounced for heart failure; for instance, the relative cumulative HGS was related to a HR of 1.92 (95%CI 1.61~2.38), which was 26.2% higher than the HR observed for HGS.

Furthermore, when categorizing HGS burdens into tertiles, a clear dose-response relationship emerged (Table S3), although not all trends were statistically significant (e.g., HGS slope’s trend *P*-value=0.166 in UKB).

### Sensitivity and subgroup analyses

In the sensitivity analyses, we found that: (i) HGS burdens or transitions remained significantly associated with incident CVD risk after excluding participants who developed CVD during the first two years after the first follow-up (Tables S4-S5) or those who underwent drug treatments at baseline and the first follow-up (Tables S6-S7). While effect estimates for HGS-related indicators remained consistent with these original results, some variations in effect sizes were observed (Figure S3). For instance, for HGS burden indicators, after excluding early incident cases, the HR of HGS slope was attenuated in some cohorts (e.g., HR=0.99 in UKB, Table S4); in contrast, the association for cumulative HGS strengthened (e.g., its HR increased from 1.65 to 2.44 in KLOSA), underscoring its role as a stable, long-term predictor of CVD risk. (ii) HGS metrics remained associated with CVD risk when accounting for competing risk of mortality (Tables S8-S9). After taking mortality into account, effect estimates for most HGS indicators increased, particularly for cumulative HGS (Figure S4), suggesting that HGS, especially cumulative burden, retains a significant link to CVD risk independent of mortality events, even though the impacts of some indicators were attenuated more in specific cohorts (e.g., HGS slope in SHARE).

Sex-stratified analyses demonstrated a 44.0% higher CVD risk increase for reduced cumulative HGS in females compared to males (HR=2.18 vs. 1.74) Tables S10-S11 and Figure S5a). Conversely, females showed a 15.4% lower risk increase for reduced relative cumulative HGS compared to males (HR=1.47 vs. 1.63), although neither difference was statistically significant (*P*=0.153 and 0.626, respectively). In the transition analysis, the harmful effect of transitioning from high to decreased HGS was more profound in females than in males (HR=1.73 vs. 1.24, Figure S6), although the correlations were not significant (*P*=0.387). Age-stratified analyses did not show pronounced changes (difference >10%) of effect sizes for burden indicators between participants aged < 65 and ≥ 65 years (Tables S12-S13 and Figure S5b).

### Improvement of model performance based on risk assessment of SCORE2

Finally, we evaluated predictive performance of the models. In European cohorts, the base model (S0, SCORE2-only) achieved an AUC ranging from 0.668 (95%CI 0.646~0.690) to 0.699 (95%CI 0.677~0.721) across the four algorithms (Figure 4a). Adding HGS and its burdens improved predictive ability, with the latter yielding higher gains. On average, compared to S1 (SCORE2+HGS), HGS slope increased the AUC by 0.3% (0.1~0.5%), cumulative HGS elevated the AUC by 1.0% (0.7~1.2%), and relative cumulative HGS created the greatest improvement of 3.8% (1.7~4.8%) across the four algorithms. The full model (S5) performed best, with improvements over S0 of 8.4% for randomForest, 7.6% for XGBoost, 7.3% for LightGBM, and 3.2% for logistic regression. LightGBM achieved the highest absolute AUC of 0.739 (95%CI 0.721~0.757), leading to an NRI of 0.217 (95%CI 0.187~0.312) and an IDI of 0.003 (95%CI 0.002~0.004) at the 7.5% risk threshold.

**Figure 4.**
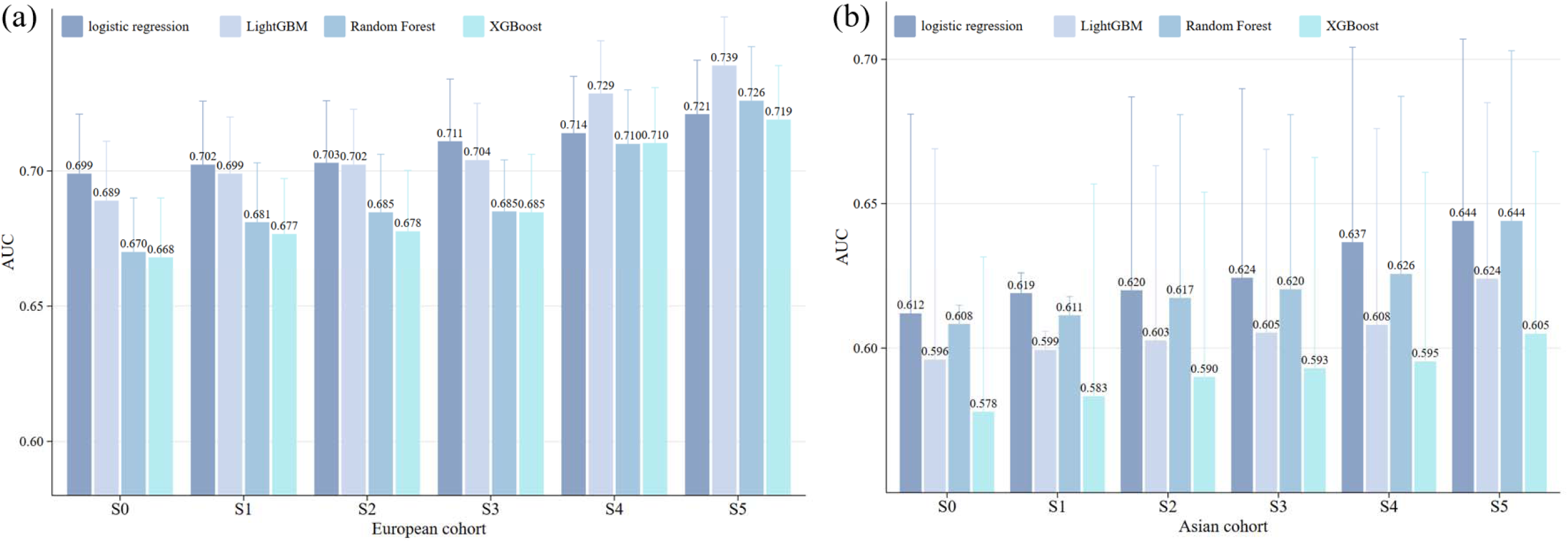
(a) Prediction accuracy in European cohort. (b) Prediction accuracy in East Asian cohort. Results of AUC for different models. Six prediction models with different sets of predictors were constructed: S0: SCORE2; S1: SCORE2 + HGS; S2: SCORE2 + HGS slope; S3: SCORE2 + cumulative HGS; S4: SCORE2 + relative cumulative HGS; S5: SCORE2 + HGS + HGS slope + cumulative HGS + relative cumulative HGS. In each model we also included available covariates, and carried out four machine learning algorithms (i.e., logistic regression, random forest, XGBoost and LightGBM), with the total samples randomly divided into training set (80%) and test set (20%).

In East Asian cohorts, S0 achieved an AUC ranging from 0.578 (95%CI 0.523~0.630) to 0.612 (95%CI 0.543~0.681) across the four algorithms (Figure 4b). Incorporating HGS (S1) into SCORE2 resulted in an average AUC increase of 0.6%, but including HGS burdens led to greater benefits. On average, compared to S1 HGS slope increased the AUC by 0.7% (0.2~1.2%), cumulative HGS by 1.2% (0.8~1.7%), and relative cumulative HGS s by 2.2% (1.5~2.9%). S5 outperformed among all settings, with enhancements over S0 by 5.9% for randomForest, 4.7% for XGBoost, 4.7% for LightGBM, and 5.2% for logistic regression. RandomForest reached the optimal AUC of 0.644, resulting in an NRI of 0.182 (95%CI 0.080~0.273) and an IDI of 0.002 (95%CI 0.001~0.003) at the 7.5% risk threshold.

We also conducted external validation to assess the model′s generalizability (Figure S7). When validated in CHARLS using models trained in European cohorts, the model performance improved consistently with the stepwise addition of HGS metrics, compatible with the internal validation trend. S5 obtained the best prediction, with AUC elevations from 0.556 (95%CI 0.515~0.597) to 0.584 (95%CI 0.559~0.609), corresponding to an improvement of 5.2~10.4% compared to S0. Similarly, the prediction models trained on CHARLS and validated in UKB or SHARE exhibited similar modest gains after adding HGS indicators. Specifically, S5 achieved an AUC increase of 11.0~19.3% in UKB and 3.1~6.2% in SHARE. These findings suggest that the predictive capability of HGS indicators for CVD risk is generally reproducible across cohorts, albeit with reduced absolute accuracy.

## Discussion

### Summary of our study

Based on four prospective cohorts, we systematically explored the association between HGS and the risk of CVD. Our findings revealed that participants whose HGS declined from high or moderate to low encountered a higher CVD risk compared to those maintaining stable HGS; in contrast, those whose HGS increased from low or moderate to higher levels exhibited a reduced risk. All HGS burden indicators were related to CVD risk in a population-specific manner. Particularly, the effect of HGS burdens was independent of and more pronounced than absolute HGS, with stronger associations observed in East Asians than in Europeans. Finally, incorporating HGS indicators into traditional CVD risk models improved predictive efficiency and enhanced the discriminative ability of existing risk scores.

### Elevation of present results and comparison with existing studies

#### Dynamic HGS trajectories and CVD risk

Overall, our findings regarding the HGS-CVD relationship are consistent with previous work demonstrating that decreased HGS increases the risk of CVD (Xie, et al., 2023). However, most prior studies relied solely on static baseline HGS; the dynamic nature of HGS, which changes over time, has remained underexplored. Additionally, existing studies mostly depended on single cohort, restricting the generalizability of their findings. By aggregating data from four cohorts totaling more than 73,000 participants, we extended cross-sectional assessments by examining longitudinal HGS trajectories and their cumulative impact on cardiovascular outcomes. While a previous study in CHARLS using absolute HGS reported that an decrease in HGS corresponded to an approximately 19% elevation in CVD risk (Zhang, et al., 2024), our analysis within the same cohort, utilizing weight-adjusted HGS, detected a much stronger effect (~59%). Moreover, we consistently observed this association across the four cohorts, greatly improving the robustness and generalizability of our findings.

Specifically, we delineated the longitudinal burden of HGS on CVD by evaluating multiple dynamic indicators, including HGS slope, cumulative HGS, and relative cumulative HGS. We discovered that declined HGS slope and lower cumulative or relative cumulative HGS were all dose-dependently associated with elevated CVD risk. Importantly, participants whose HGS increased or kept stable over time had better cardiovascular health, underscoring the potential reversibility of risk through muscular health maintenance. A recent analysis within the UKB alone similarly demonstrated that individuals with declining or sustained low HGS trajectories faced an elevated risk of incident CVD, and this longitudinal association remained significant even after adjusting for frailty phenotypes. Furthermore, their findings showed that an accelerated annual rate of HGS decline continuously increased CVD risk, which strongly corroborates our results regarding the dose-dependent harm of a declining HGS slope (Chai, et al., 2026). However, this study only provides crucial dynamic insights within a single European cohort, while our study significantly extends this framework. Notably, by introducing more comprehensive long-term exposure metrics, specifically cumulative and relative cumulative HGS, and systematically validating these dynamic relationships across four diverse cohorts, our findings provide deeper insights into the cross-ethnic consistency and variations of HGS burdens.

#### Racial differences in HGS burdens

Although existing studies have not yet clarified whether there are significant differences in the association between HGS burdens and CVD among different ethnic groups, previous cross-ethnic comparisons regarding HGS and diabetes have suggested that racial factors play an important role in the relationship between HGS and health outcomes (Qiu, et al., 2023). We discovered that the effects of HGS burdens varied substantially across cohorts, and were more pronounced in East Asian cohorts, highlighting the importance of conducting HGS-related cardiovascular risk assessments in a multi-ethnic context.

This discrepancy may be partly related to differences in body composition among races. Compared to Europeans, East Asians generally have lower baseline muscle mass (Ntuk, et al., 2017), and HGS is a reliable indicator of overall muscle function. In groups with lower baseline muscle mass, a decline in HGS may indicate the occurrence and progression of sarcopenia more quickly (Haldar, et al., 2015; Rush, et al., 2009). Notably, East Asians frequently exhibit a distinct phenotype of sarcopenic obesity, characterized by diminished skeletal muscle mass and suboptimal gait speed despite a relatively low BMI. This profile is often accompanied by a disproportionately high accumulation of visceral adiposity. Such a metabolic configuration implies that even a marginal decline in HGS, acting as a proxy for compromised muscle quality, could potently amplify the deleterious effects of hidden obesity (Axelrod, et al., 2023; Chen, et al., 2025). Mechanistically, the synergy between visceral fat accrual and muscle depletion precipitates systemic chronic low-grade inflammation and oxidative stress, both of which are critical drivers of vascular endothelial impairment. Furthermore, the accumulation of intramyocellular lipids, also known as myosteatosis, induces lipotoxicity and anabolic resistance (Amartuvshin, et al., 2020; Fealy, et al., 2018). When these factors are superimposed on the aging process, they severely undermine muscle strength and physical performance (Cruz-Jentoft and Sayer, 2019). Therefore, the same magnitude of the HGS burden decline in East Asians may indicate a more severe state of muscle loss, consequently leading to a higher cardiovascular risk (Li, et al., 2024). In addition, prior studies have indicated that inter-ethnic differences in body composition may influence the relationship between muscle function and metabolic health, further modulating the association between HGS burdens and CVD risk (Abe, et al., 2012).

#### Added predictive performance

We utilized distinct machine learning models to evaluate the predictive value of both static HGS and its dynamic burden indicators for CVD risk. In both European and East Asian cohorts, no single algorithm was universally superior across all scenarios. Additionally, the prediction accuracy in East Asian cohorts was typically lower than that in European cohorts, likely due to smaller sample sizes. Among all considered settings, the full LightGBM model demonstrated the highest accuracy in European cohorts, achieving a 7.6% increase in AUC compared to SCORE2. In East Asian cohorts, the full randomForest model performed best, yielding a 5.9% increase in AUC over SCORE2. The relative cumulative HGS emerged as the single most influential dynamic indicator in both populations, contributing the largest performance improvement among three burden metrics when adding them individually. This finding demonstrates that integrating HGS, particularly the cumulative burden indicator which captures long-term exposure, into established risk models can enhance the prediction of CVD risk across diverse ethnic populations.

#### Advantages of burden indicators over a single cross-sectional HGS

We consistently observed that, compared to a single cross-sectional HGS measurement, these burden indicators were more strongly associated with CVD risk and offered greater predictive improvement. Several reasons may explain this phenomenon. First, a single HGS measurement provides only a static snapshot of muscle strength at a single time point, which fails to adequately reflect the cumulative physiological stress or adaptive capacity of the musculoskeletal and cardiovascular systems over time (Han, et al., 2025). In contrast, burden metrics integrate multiple longitudinal measurements, thereby capturing the trajectory of muscle strength change (Ling, et al., 2010; López-Bueno, et al., 2022).

Second, HGS burden indicators are typically less susceptible to measurement variability and transient fluctuations. A single HGS measurement can be influenced by acute factors such as recent physical activity, fatigue, or minor hand discomfort (Macdonald, et al., 2022). By integrating measurements from multiple time points, HGS burden metrics reduce the impact of such variability and offer a more stable estimate of an individual’s underlying muscle function trajectory.

Third, the decline in HGS is not a transient event but is closely related to processes such as chronic inflammation and metabolic disorders, which have cumulative, time-dependent effects (Shen, et al., 2020). A single HGS measurement cannot capture this dynamic evolution, whereas the HGS burden indices can successfully integrate the time dimension and reflect the long-term state of muscle metabolic function and the sustained load on the cardiovascular system more directly (Yi, et al., 2018). For instance, HGS slope reflects the decline speed of muscle function, which is closer to the pathological process of CVD than the static HGS. Cumulative HGS reflects the continuous health exposure level by integrating long-term muscle function reserve; relative cumulative HGS quantifies dynamic exposure deviation from baseline, which is a potential direct manifestation of metabolic disorder and vascular injury.

### Public health implications

Our findings carry significant public health implications. First, establishing the dynamic trajectory of HGS as a key, modifiable determinant of cardiovascular health provides an empirical foundation for integrating muscle strength monitoring into public health frameworks. The observed reduction in cardiovascular risk among individuals maintaining stable or improved HGS trajectories underscores the preventive potential of targeted interventions (Zhang, et al., 2024).

Second, dynamic HGS monitoring, as a simple and low-cost indicator of muscle function, represents an effective tool for screening individuals at elevated cardiovascular risk in primary care settings. Our results confirm that lower HGS significantly elevates CVD risk, while higher HGS is protective, lending support to promoting regular, community-based HGS monitoring for early identification of at-risk groups (Willis, et al., 2012). Both cumulative and relative cumulative HGS demonstrate superior predictive value for CVD risk compared to single time-point assessments, indicating that public health interventions should prioritize long-term muscle function trajectories over solely cross-sectional measurements.

Third, our findings indirectly imply that the biological aging process, as reflected by HGS and its burdens (Kemala Sari, et al., 2025; Vaishya, et al., 2024; Vennu, 2023), is not entirely irreversible. The fact that an improvement in a modifiable marker like HGS is associated with a lower risk of CVD supports the concept that the pace of biological aging can be slowed. By identifying HGS as a tractable indicator and potential interventional target, our study bolsters the rationale for mitigating age-related health risks through targeted lifestyle modifications and medical interventions.

Finally, integrating HGS metrics into the established SCORE2 risk assessment model enhanced its predictive efficacy. This suggests that incorporating dynamic HGS assessment into existing CVD prevention frameworks could enhance risk stratification cost-effectively, facilitating a more efficient allocation of healthcare resources.

### Limitations of our study

Several limitations should be mentioned. First, although standardized HGS measurements were used, they may still be affected by individual differences such as hand-specific conditions or recent activity levels, which could influence their predictive accuracy (Celis-Morales, et al., 2018; Zhang, et al., 2024). Second, although we conducted sensitivity analyses by excluding participants undergoing specific drug treatments, our primary models did not fully adjust for the impacts of all medication use or specific comorbidities. This residual confounding may affect the observed associations between HGS burdens and CVD risk. Third, the heterogeneity of CVD definitions across cohorts likely influenced the comparability of results. Specifically, CVD was defined based on ICD codes in the UK Biobank but relied on self-reported stroke and heart problems in other cohorts, leading to potential heterogeneity in outcome ascertainment.

## Conclusions

This study identifies dynamic HGS trajectories and cumulative burdens as key predictors of cardiovascular health. The inclusion of these indices demonstrates moderate yet consistent incremental value over traditional cardiovascular risk prediction models. These findings support the incorporation of longitudinal HGS monitoring into clinical and public health strategies to reduce the global burden of CVD. However, future research should focus on underlying biological mechanisms and interventions targeting HGS preservation as a modifiable risk factor.

## Additional File

Supplementary File

## Abbreviations

CVD: cardiovascular disease
HGS: handgrip strength
BMI: body mass index
SD: standard deviation
HR: hazard ratio
CI: confidence interval
CHARLS: China Health and Retirement Longitudinal Study
UKB: UK Biobank
SHARE: Survey of Health, Ageing and Retirement in Europe
KLOSA: Korean Longitudinal Study of Ageing
MICE: multivariate imputation by chained equations
NRI: net reclassification improvement
IDI: integrated discrimination improvement
IQR: interquartile range
AUC: Area under the curve

## Acknowledgments

This study was partly based on the UK Biobank resource under application number 88159. The UK Biobank was established by the Wellcome Trust medical charity, Medical Research Council, Department of Health, Scottish Government, and the Northwest Regional Development Agency. It has also had funding from the Welsh Assembly Government, British Heart Foundation and Diabetes UK. This study also relied on CHARLS, SHARE and KLOSA, the authors express their great gratitude to these research teams and all individuals who participated in the studies. The data analyses in the present study were carried out with the high-performance computing cluster that was supported by the special central finance project of local universities for Xuzhou Medical University.

## Funding

This research was supported in part by the Project of Philosophy and Social Science Research in Colleges and Universities of Jiangsu Province (2024SJYB0809) and the Youth Foundation of Humanity and Social Science funded by the Ministry of Education of China (18YJC910002).

## Contributors

PZ conceived the idea for the study. XH obtained the data. PZ and LH, JK cleared up the datasets; HL performed the data analyses. PZ, KW, HL, JQ, YJ, YY, XZ, JS, MS, TW, WC, CW, TX, TX and CZ interpreted the results of the data analyses. PZ and HL wrote the manuscript with the help from other authors.

## Data availability

All data generated or analyzed during this study are included in this published article and its supplementary information files. This study used the UK Biobank resource with the application ID 88159. Researchers can access to the UK Biobank by applying to the UK Biobank official website (https://www.ukbiobank.ac.uk/). The CHARLS, SHARE and KLOSA data were publicly available from http://charls.pku.edu.cn/, https://releases.sharedataportal.eu/, and https://survey.keis.or.kr/eng/index.jsp, respectively.

## Ethics approval and consent to participate

The original surveys granted ethical approval and were conducted in accordance with the principles of the Declaration of Helsinki. Informed consent was obtained from all participants using the original surveys.

## Consent for publication

All authors have approved the manuscript and given their consent for submission and publication.

## Competing interests

The authors declare that they have no competing interests.

## Notes

### Competing Interest Statement

The authors have declared no competing interest.

### Author Declarations

The datasets used in your study (CHARLS, SHARE and KLOSA) were individual-level data, but any individual-level data had been de-identified prior to use in this study.

## References

1 Abe, T., Bemben, M.G., Kondo, M., et al. (2012). Comparison of skeletal muscle mass to fat-free mass ratios among different ethnic groups, The journal of nutrition, health & aging, 16, 534–538.

2 Amartuvshin, O., Lin, C.H., Hsu, S.C., et al. (2020). Aging shifts mitochondrial dynamics toward fission to promote germline stem cell loss, Aging cell, 19, e13191.

3 Andersen-Ranberg, K., Petersen, I., Frederiksen, H., et al. (2009). Cross-national differences in grip strength among 50+ year-old Europeans: results from the SHARE study, Eur J Ageing, 6, 227–236.

4 Andersen, K., Rasmussen, F., Held, C., et al. (2015). Exercise capacity and muscle strength and risk of vascular disease and arrhythmia in 1.1 million young Swedish men: cohort study, BMJ, 351, h4543.

5 Axelrod, C.L., Dantas, W.S. and Kirwan, J.P. (2023). Sarcopenic obesity: emerging mechanisms and therapeutic potential, Metabolism, 146, 155639.

6 Bae, K.H., Jo, Y.H., Lee, D.R., et al. (2021). Trajectories of Handgrip Strength and Their Associations with Mortality among Older Adults in Korea: Analysis of the Korean Longitudinal Study of Aging, Korean journal of family medicine, 42, 38–46.

7 Bhat, A.K., Jindal, R. and Acharya, A.M. (2021). The influence of ethnic differences based on upper limb anthropometry on grip and pinch strength, Journal of clinical orthopaedics and trauma, 21, 101504.

8 Celis-Morales, C.A., Welsh, P., Lyall, D.M., et al. (2018). Associations of grip strength with cardiovascular, respiratory, and cancer outcomes and all cause mortality: prospective cohort study of half a million UK Biobank participants, BMJ, 361, k1651.

9 Cesari, M., Penninx, B.W., Pahor, M., et al. (2004). Inflammatory markers and physical performance in older persons: the InCHIANTI study, The journals of gerontology. Series A, Biological sciences and medical sciences, 59, 242–248.

10 Chai, L., Zhang, Y., Zhang, K., et al. (2026). Association of changes in grip strength with risk of incident cardiovascular disease in middle aged and older adults from the UK biobank, BMC public health.

11 Chen, T. and Guestrin, C. XGBoost: A Scalable Tree Boosting System. In, Proceedings of the 22nd ACM SIGKDD International Conference on Knowledge Discovery and Data Mining. San Francisco, California, USA: Association for Computing Machinery; 2016. p. 785–794.

12 Chen, T.P., Kao, H.H., Ogawa, W., et al. (2025). The Asia-Oceania consensus: Definitions and diagnostic criteria for sarcopenic obesity, Obes Res Clin Pract, 19, 185–192.

13 Cheung, C.L., Tan, K.C., Bow, C.H., et al. (2012). Low handgrip strength is a predictor of osteoporotic fractures: cross-sectional and prospective evidence from the Hong Kong Osteoporosis Study, Age (Dordr), 34, 1239–1248.

14 Chi, J.H. and Lee, B.J. (2024). Association of relative hand grip strength with myocardial infarction and angina pectoris in the Korean population: a large-scale cross-sectional study, BMC public health, 24, 941.

15 Choquette, S., Bouchard, D.R., Doyon, C.Y., et al. (2010). Relative strength as a determinant of mobility in elders 67-84 years of age. a nuage study: nutrition as a determinant of successful aging, The journal of nutrition, health & aging, 14, 190–195.

16 Cruz-Jentoft, A.J. and Sayer, A.A. (2019). Sarcopenia, Lancet, 393, 2636–2646.

17 Cui, H., Liu, Q., Wu, Y., et al. (2022). Cumulative triglyceride-glucose index is a risk for CVD: a prospective cohort study, Cardiovasc Diabetol, 21, 22.

18 D’Agostino, R.B., Sr., Vasan, R.S., Pencina, M.J., et al. (2008). General cardiovascular risk profile for use in primary care: the Framingham Heart Study, Circulation, 117, 743–753.

19 Ding, N., Ballew, S.H., Palta, P., et al. (2020). Muscle Strength and Incident Cardiovascular Outcomes in Older Adults, Journal of the American College of Cardiology, 75, 1090–1092.

20 Dodds, R.M., Syddall, H.E., Cooper, R., et al. (2016). Global variation in grip strength: a systematic review and meta-analysis of normative data, Age and ageing, 45, 209–216.

21 Dong, R., Wang, X., Guo, Q., et al. (2016). Clinical Relevance of Different Handgrip Strength Indexes and Mobility Limitation in the Elderly Adults, The journals of gerontology. Series A, Biological sciences and medical sciences, 71, 96–102.

22 Duchowny, K.A., Ackley, S.F., Brenowitz, W.D., et al. (2022). Associations Between Handgrip Strength and Dementia Risk, Cognition, and Neuroimaging Outcomes in the UK Biobank Cohort Study, JAMA Network Open, 5, e2218314.

23 Fealy, C.E., Mulya, A., Axelrod, C.L., et al. (2018). Mitochondrial dynamics in skeletal muscle insulin resistance and type 2 diabetes, Transl Res, 202, 69–82.

24 Fine, J.P. and Gray, R.J. (1999). A Proportional Hazards Model for the Subdistribution of a Competing Risk, Journal of the American Statistical Association, 94, 496–509.

25 Gao, K., Cao, L.F., Ma, W.Z., et al. (2022). Association between sarcopenia and cardiovascular disease among middle-aged and older adults: Findings from the China health and retirement longitudinal study, EClinicalMedicine, 44, 101264.

26 Grundy, S.M., Stone, N.J., Bailey, A.L., et al. (2019). 2018 AHA/ACC/AACVPR/AAPA/ABC/ACPM/ADA/AGS/APhA/ASPC/NLA/PC NA Guideline on the Management of Blood Cholesterol: A Report of the American College of Cardiology/American Heart Association Task Force on Clinical Practice Guidelines, Circulation, 139, e1082–e1143.

27 Haldar, S., Chia, S.C. and Henry, C.J. (2015). Body Composition in Asians and Caucasians: Comparative Analyses and Influences on Cardiometabolic Outcomes, Advances in food and nutrition research, 75, 97–154.

28 Han, B., Zeng, Z., Wen, Y., et al. (2025). Cumulative handgrip strength and longitudinal changes in cognitive function and daily functioning among people aged 50 years and older: evidence from two longitudinal cohort studies, Arch Public Health, 83, 150.

29 Hardy, R., Cooper, R., Aihie Sayer, A., et al. (2013). Body mass index, muscle strength and physical performance in older adults from eight cohort studies: the HALCyon programme, PloS one, 8, e56483.

30 He, D., Wang, Z., Li, J., et al. (2024). Changes in frailty and incident cardiovascular disease in three prospective cohorts, European heart journal, 45, 1058–1068.

31 Ho, F.K.W., Celis-Morales, C.A., Petermann-Rocha, F., et al. (2019). The association of grip strength with health outcomes does not differ if grip strength is used in absolute or relative terms: a prospective cohort study, Age and ageing, 48, 684–691.

32 Jiang, R., Westwater, M.L., Noble, S., et al. (2022). Associations between grip strength, brain structure, and mental health in > 40,000 participants from the UK Biobank, BMC Med., 20, 286.

33 Kang, K.Y., Jung, Y.E., Jang, H., et al. (2021). Relationship between Handgrip Strength and Low-grade Inflammation in Older Adults with Depression, Clin Psychopharmacol Neurosci, 19, 721–730.

34 Ke, G., Meng, Q., Finley, T., et al. LightGBM: A Highly Efficient Gradient Boosting Decision Tree. In, Neural Information Processing Systems. 2017.

35 Kemala Sari, N., Stepvia, S., Ilyas, M.F., et al. (2025). Handgrip strength as a potential indicator of aging: insights from its association with aging-related laboratory parameters, Frontiers in medicine, 12, 1491584.

36 Kim, Y.M., Kim, S., Bae, J., et al. (2021). Association between relative hand-grip strength and chronic cardiometabolic and musculoskeletal diseases in Koreans: A cross-sectional study, Archives of gerontology and geriatrics, 92, 104181.

37 Kothari, R., Johny, M.P., Mistry, D., et al. (2024). Grip to Health: Unlocking the Clinical Potential of Isometric Hand Grip Strength - A Narrative Review, Journal of pharmacy & bioallied sciences, 16, s3102–s3104.

38 Kuo, K., Zhang, Y.-R., Chen, S.-D., et al. (2023). Associations of grip strength, walking pace, and the risk of incident dementia: A prospective cohort study of 340212 participants, Alzheimer’s & Dementia, 19, 1415–1427.

39 Laukkanen, J.A., Voutilainen, A., Kurl, S., et al. (2020). Handgrip strength is inversely associated with fatal cardiovascular and all-cause mortality events, Annals of medicine, 52, 109–119.

40 Lawman, H.G., Troiano, R.P., Perna, F.M., et al. (2016). Associations of Relative Handgrip Strength and Cardiovascular Disease Biomarkers in U.S. Adults, 2011-2012, American journal of preventive medicine, 50, 677–683.

41 Lee, W.J., Peng, L.N., Chiou, S.T., et al. (2016). Relative Handgrip Strength Is a Simple Indicator of Cardiometabolic Risk among Middle-Aged and Older People: A Nationwide Population-Based Study in Taiwan, PloS one, 11, e0160876.

42 Leong, D.P., Teo, K.K., Rangarajan, S., et al. (2015). Prognostic value of grip strength: findings from the Prospective Urban Rural Epidemiology (PURE) study, Lancet, 386, 266–273.

43 Li, D., Guo, G., Xia, L., et al. (2018). Relative Handgrip Strength Is Inversely Associated with Metabolic Profile and Metabolic Disease in the General Population in China, Frontiers in physiology, 9, 59.

44 Li, G., Lu, Y., Shao, L., et al. (2023). Handgrip strength is associated with risks of new-onset stroke and heart disease: results from 3 prospective cohorts, BMC geriatrics, 23, 268.

45 Li, H., Zheng, Y., Zhang, Y., et al. (2024). Handgrip strength and body mass index exhibit good predictive value for sarcopenia in patients on peritoneal dialysis, Frontiers in nutrition, 11, 1470669.

46 Liaw, A. and Wiener, M.C. Classification and Regression by randomForest. In.; 2007.

47 Ling, C.H., Taekema, D., de Craen, A.J., et al. (2010). Handgrip strength and mortality in the oldest old population: the Leiden 85-plus study, CMAJ, 182, 429–435.

48 Liu, W., Chen, R., Song, C., et al. (2021). A Prospective Study of Grip Strength Trajectories and Incident Cardiovascular Disease, Frontiers in cardiovascular medicine, 8, 705831.

49 López-Bueno, R., Andersen, L.L., Calatayud, J., et al. (2022). Longitudinal association of handgrip strength with all-cause and cardiovascular mortality in older adults using a causal framework, Exp Gerontol, 168, 111951.

50 López-Bueno, R., Andersen, L.L., Koyanagi, A., et al. (2022). Thresholds of handgrip strength for all-cause, cancer, and cardiovascular mortality: A systematic review with dose-response meta-analysis, Ageing research reviews, 82, 101778.

51 Macdonald, G.A., Manning, J.W., Bodell, N.G., et al. (2022). Acute Handgrip Fatigue and Forearm Girth in Recreational Sport Rock Climbers, Int J Exerc Sci, 15, 834–845.

52 Magnussen, C., Ojeda, F.M., Leong, D.P., et al. (2023). Global Effect of Modifiable Risk Factors on Cardiovascular Disease and Mortality, The New England journal of medicine, 389, 1273–1285.

53 Mainous, A.G., 3rd, Tanner, R.J., Anton, S.D., et al. (2015). Grip Strength as a Marker of Hypertension and Diabetes in Healthy Weight Adults, American journal of preventive medicine, 49, 850–858.

54 Marcotte-Chénard, A., Oliveira, B., Little, J.P., et al. (2023). Sarcopenia and type 2 diabetes: Pathophysiology and potential therapeutic lifestyle interventions, Diabetes & metabolic syndrome, 17, 102835.

55 Min, J.Y., Lee, K.J., Park, J.B., et al. (2012). Social engagement, health, and changes in occupational status: analysis of the Korean Longitudinal Study of Ageing (KLoSA), PloS one, 7, e46500.

56 Ntuk, U.E., Celis-Morales, C.A., Mackay, D.F., et al. (2017). Association between grip strength and diabetes prevalence in black, South-Asian, and white European ethnic groups: a cross-sectional analysis of 418656 participants in the UK Biobank study, Diabet Med, 34, 1120–1128.

57 Peterson, M.D., Zhang, P., Choksi, P., et al. (2016). Muscle Weakness Thresholds for Prediction of Diabetes in Adults, Sports Med, 46, 619–628.

58 Prasitsiriphon, O. and Pothisiri, W. (2018). Associations of Grip Strength and Change in Grip Strength With All-Cause and Cardiovascular Mortality in a European Older Population, Clinical Medicine Insights. Cardiology, 12, 1179546818771894.

59 Qiu, S., Wang, Q., Chen, W., et al. (2023). Cumulative Muscle Strength and Risk of Cardiovascular Disease and All-cause mortality: A Prospective Cohort Study, Archives of medical research, 54, 261–269.

60 Roth, G.A., Mensah, G.A., Johnson, C.O., et al. (2020). Global Burden of Cardiovascular Diseases and Risk Factors, 1990-2019: Update From the GBD 2019 Study, Journal of the American College of Cardiology, 76, 2982–3021.

61 Rush, E.C., Freitas, I. and Plank, L.D. (2009). Body size, body composition and fat distribution: comparative analysis of European, Maori, Pacific Island and Asian Indian adults, The British journal of nutrition, 102, 632–641.

62 Samarasekera, E.J., Clark, C.E., Kaur, S., et al. (2023). Cardiovascular disease risk assessment and reduction: summary of updated NICE guidance, BMJ, 381, 1028.

63 SCORE2 working group and ESC Cardiovascular risk collaboration (2021). SCORE2 risk prediction algorithms: new models to estimate 10-year risk of cardiovascular disease in Europe, European heart journal, 42, 2439–2454.

64 Shen, C., Lu, J., Xu, Z., et al. (2020). Association between handgrip strength and the risk of new-onset metabolic syndrome: a population-based cohort study, BMJ open, 10, e041384.

65 Studenski, S.A., Peters, K.W., Alley, D.E., et al. (2014). The FNIH sarcopenia project: rationale, study description, conference recommendations, and final estimates, The journals of gerontology. Series A, Biological sciences and medical sciences, 69, 547–558.

66 Sudlow, C., Gallacher, J., Allen, N., et al. (2015). UK biobank: an open access resource for identifying the causes of a wide range of complex diseases of middle and old age, PLoS medicine, 12, e1001779.

67 Sun, Y., Li, W., Zhou, Y., et al. (2024). Long-term changes in frailty and incident type 2 diabetes: A prospective cohort study based on the UK Biobank, Diabetes Obes Metab, 26, 3352–3360.

68 Sun, Y., Sun, M., Zeng, X., et al. (2025). Effects of resistance training on cardiovascular risk factors in patients with type 2 diabetes mellitus: a systematic review and meta-analysis of randomized controlled trials, Acta diabetologica, 62, 11–24.

69 Tikkanen, E., Gustafsson, S. and Ingelsson, E. (2018). Associations of Fitness, Physical Activity, Strength, and Genetic Risk With Cardiovascular Disease: Longitudinal Analyses in the UK Biobank Study, Circulation, 137, 2583–2591.

70 Vaishya, R., Misra, A., Vaish, A., et al. (2024). Hand grip strength as a proposed new vital sign of health: a narrative review of evidences, Journal of health, population, and nutrition, 43, 7.

71 van Buuren, S. and Groothuis-Oudshoorn, K. (2011). mice: Multivariate Imputation by Chained Equations in R, Journal of Statistical Software, 45, 1–67.

72 van der Kooi, A.L., Snijder, M.B., Peters, R.J., et al. (2015). The Association of Handgrip Strength and Type 2 Diabetes Mellitus in Six Ethnic Groups: An Analysis of the HELIUS Study, PloS one, 10, e0137739.

73 Vennu, V. (2023). Biological ageing and the risk of decreased handgrip strength among community-dwelling older adult Indians: a cross-sectional study, BMC geriatrics, 23, 782.

74 Wen, Y., Liu, T., Ma, C., et al. (2022). Association between handgrip strength and metabolic syndrome: A meta-analysis and systematic review, Frontiers in nutrition, 9, 996645.

75 Willis, A., Davies, M., Yates, T., et al. (2012). Primary prevention of cardiovascular disease using validated risk scores: a systematic review, Journal of the Royal Society of Medicine, 105, 348–356.

76 Wu, H., Wang, D., Wang, X., et al. (2025). The association between handgrip strength and metabolic syndrome: A large prospective Chinese cohort study, Maturitas, 192, 108157.

77 Xiang, Y., Xu, H., Chen, H., et al. (2024). Tea consumption and attenuation of biological aging: a longitudinal analysis from two cohort studies, Lancet Reg Health West Pac, 42, 100955.

78 Xie, K.H., Han, X., Zheng, W.J., et al. (2023). Low Grip Strength and Increased Mortality Hazard among Middle-Aged and Older Chinese Adults with Chronic Diseases, Biomed Environ Sci, 36, 213–221.

79 Xie, Z., Wang, L., Sun, M., et al. (2023). Mediation of 10-Year Cardiovascular Disease Risk between Inflammatory Diet and Handgrip Strength: Base on NHANES 2011-2014, Nutrients, 15, 918.

80 Yang, Z., Wei, J., Liu, H., et al. (2024). Changes in muscle strength and risk of cardiovascular disease among middle-aged and older adults in China: Evidence from a prospective cohort study, Chinese medical journal, 137, 1343–1350.

81 Yi, D.W., Khang, A.R., Lee, H.W., et al. (2018). Relative handgrip strength as a marker of metabolic syndrome: the Korea National Health and Nutrition Examination Survey (KNHANES) VI (2014-2015), Diabetes Metab Syndr Obes, 11, 227–240.

82 Yu, Y., Tang, Y., Li, X., et al. (2025). Association of physical activity, cardiorespiratory fitness, grip strength, and grip strength asymmetry with incident musculoskeletal disorders in 406,080 White adults, Journal of Sport and Health Science, 14, 101040.

83 Zammit, A.R., Piccinin, A.M., Duggan, E.C., et al. (2021). A Coordinated Multi-study Analysis of the Longitudinal Association Between Handgrip Strength and Cognitive Function in Older Adults, J Gerontol B Psychol Sci Soc Sci, 76, 229–241.

84 Zeng, P. and Zhou, X. (2017). Non-parametric genetic prediction of complex traits with latent Dirichlet process regression models, Nature communications, 8, 456.

85 Zhang, F., Luo, B., Bai, Y., et al. (2024). Association of handgrip strength and risk of cardiovascular disease: a population-based cohort study, Aging clinical and experimental research, 36, 207.

86 Zhang, X., Yan, Y., Liu, Y., et al. (2025). Association of biological aging acceleration transitions and burdens with incident cardiovascular disease: longitudinal insights from a national cohort study, BMC Med, 23, 347.

87 Zhang, X.L., Gu, Y., Zhao, J., et al. (2025). Associations between skeletal muscle strength and chronic kidney disease in patients with MASLD, Communications medicine, 5, 118.

88 Zhao, D., Wang, Y., Wong, N.D., et al. (2024). Impact of Aging on Cardiovascular Diseases, JACC: Asia, 4, 345–358.

89 Zhao, Y., Hu, Y., Smith, J.P., et al. (2014). Cohort profile: the China Health and Retirement Longitudinal Study (CHARLS), International journal of epidemiology, 43, 61–68.

