## Supplemental Table 1 for "Cross-cohort insights into the association of handgrip strength transitions and burdens with cardiovascular disease risk"

**Supplementary File**

### Table S1. Definitions and sources of CVD in the UK Biobank.

| Disease | | Field | Description of field | Codes |
| --- | --- | --- | --- | --- |
| CVD | CHD | 41270 | ICD-10 | I20-I25, Z951, Z955 |
|  |  | 41271 | ICD-9 | 410-414 |
|  | Atrial fibrillation | 41270 | ICD-10 | I48 |
|  |  | 41271 | ICD-9 | 4273 |
|  | Heart failure | 41270 | ICD-10 | I50 |
|  |  | 41271 | ICD-9 | 428 |
|  | Stroke | 41270 | ICD-10 | I60, I61, I629, I63, I64, I678, I690, I693 |
|  |  | 41271 | ICD-9 | 430, 431, 434, 436, 3361, 3623, 4329, 4331, 4332, 4333, 4338, 4339 |
|  |  | 42006 | Algorithmically defined | - |
|  |  | 42008 |  | - |
|  |  | 42010 |  | - |
|  |  | 42012 |  | - |
|  | Myocardial infarction | 41270 | ICD-10 | I21, I22, I23, I241, I252 |
|  |  | 41271 | ICD-9 | 410, 411, 412, 429 |

Note: CVD: cardiovascular disease; CHD: coronary heart disease; ICD: International Classification of Disease.

### Table S2. Calculation of the SCORE2 algorithm.

| Risk factor (units) | Transformation equation | Log sHR | | Calculation of 10-year cumulative incidence |
| --- | --- | --- | --- | --- |
|  |  | Male | Female |  |
| Age (years) | cage = (age - 60)/5 | 0.3742 | 0.4648 | **Male:**  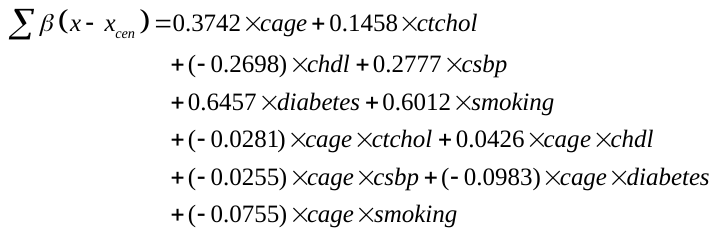  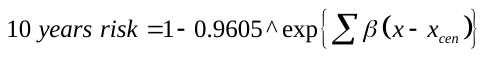  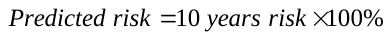  **Female:**  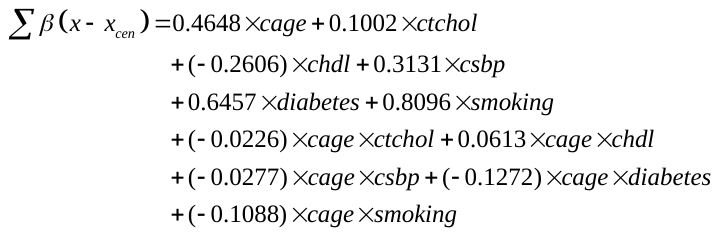  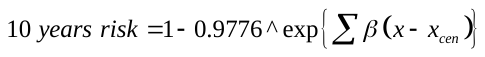  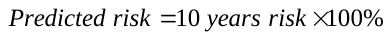 |
| Smoking (current or others) | yes = 1, no = 1 | 0.6012 | 0.7744 |  |
| Diabetes (yes or no) | yes = 1, no = 1 | 0.6457 | 0.8096 |  |
| SBP (mmHg) | csbp = (SBP – 120)/20 | 0.2777 | 0.3131 |  |
| Total cholesterol (mmol/L) | ctchol = (tchol – 6)/1 | 0.1458 | 0.1002 |  |
| HDL cholesterol (mg/dL) | chdl = (hdl -1.3)/0.5d | -0.2698 | -0.2606 |  |
| Smoking × age interaction | cage × smoking | -0.0755 | -0.1088 |  |
| SBP × age interaction | cage × csbp | -0.0255 | -0.0277 |  |
| Total cholesterol × age interaction | cage × ctchol | -0.0281 | -0.0226 |  |
| HDL cholesterol × age interaction | cage × chdl | 0.0426 | 0.0613 |  |
| Diabetes ×age interaction | cage × diabetes | -0.0983 | -0.1272 |  |

Note: SBP: systolic blood pressure; HDL: high-density lipoprotein; sHR: standardized hazard ratio. Total cholesterol and HDL cholesterol were lacked in SHARE, and their corresponding coefficients were set to zero when SCORE2 was calculated.

### Table S3. Association of HGS burdens with incident CVD risk.

| Burdens | | UKB | | | CHARLS | | | SHARE | | | KLOSA | | |
| --- | --- | --- | --- | --- | --- | --- | --- | --- | --- | --- | --- | --- | --- |
|  |  | Events  (cases/non-cases) | HR  (95%CI) | *P*-value | Events  (cases/non-cases) | HR  (95%CI) | *P*-value | Events  (cases/non-cases) | HR  (95%CI) | *P*-value | Events  (cases/non-cases) | HR  (95%CI) | *P*-value |
| Slope | Continuous | 8,698/37,198 | 0.99 (0.96,1.03) | 0.061 | 478/5,149 | 1.33 (1.12,1.57) | 2.19×10^-8^ | 951/23,277 | 1.11 (1.04,1.19) | 3.01×10^-3^ | 278/4,335 | 1.43 (1.35,1.51) | 3.82×10^-8^ |
|  | T1 | 2,671/12,054 | 1.00 (Ref.) | - | 154/1,645 | 1.00 (Ref.) | - | 281/7,451 | 1.00 (Ref.) | - | 81/1,324 | 1.00 (Ref.) | - |
|  | T2 | 2,812/12,217 | 1.02 (0.94,1.11) | 0.591 | 155/1,732 | 1.41 (1.14,1.75) | 2.21×10^-5^ | 325/7,781 | 1.15 (1.03,1.27) | 8.39×10^-5^ | 98/1,471 | 1.20 (0.98,1.46) | 0.105 |
|  | T3 | 3,215/12,927 | 1.14 (1.06,1.22) | 0.105 | 169/1,772 | 1.72 (1.37,2.22) | 8.87×10^-6^ | 345/8.045 | 1.22 (1.06,1.37) | 1.30×10^-4^ | 99/1,540 | 2.13 (1.49,2.78) | 1.97×10^-5^ |
|  | *P* for trend | 0.166 | | | 2.67×10^-3^ | | | 1.19×10^-4^ | | | 4.12×10^-5^ | | |
| Cumulative | Continuous | 8,698/37,198 | 1.05 (1.02,1.09) | 3.76×10^-6^ | 478/5,149 | 1.89 (1.54,2.33) | 1.72×10^-3^ | 951/23,277 | 1.39 (1.12,1.72) | 5.12×10^-3^ | 278/4,335 | 1 65 (1.33,2.06) | 2.71×10^-7^ |
|  | T1 | 2,712/11,232 | 1.00 (Ref.) | - | 129/1,527 | 1.00 (Ref.) | - | 298/7,712 | 1.00 (Ref.) | - | 87/1,254 | 1.00 (Ref.) | - |
|  | T2 | 2,854/12,205 | 1.16 (1.10,1.25) | 1.04×10^-6^ | 165/1,741 | 1.59 (1.29,1.96) | 7.81×10^-6^ | 312/7,584 | 1.37 (1.12,1.67) | 2.01×10^-5^ | 94/1,312 | 1.56 (1.09,2.27) | 1.87×10^-3^ |
|  | T3 | 3,132/13,761 | 1.25 (1.15,1.35) | 2.81×10^-7^ | 184/1,881 | 1.82 (1.54,2.10) | 1.88×10^-5^ | 341/7,981 | 1.49 (1.11,1.84) | 9.53×10^-5^ | 97/1,769 | 2.38 (1.79,2.94) | 7.37×10^-5^ |
|  | *P* for trend | 3.17×10^-7^ | | | 5.19×10^-5^ | | | 9.32×10^-5^ | | | 2.19×10^-4^ | | |
| Relative  cumulative | Continuous | 8,698/37,198 | 1.11 (1.06,1.15) | 3.62×10^-6^ | 478/5,149 | 1.43 (1.27,1.59) | 4.12×10^-3^ | 951/23,277 | 1.14 (1.06,1.20) | 2.12×10^-2^ | 278/4,335 | 1.49 (1.28,1.72) | 8.32×10^-5^ |
|  | T1 | 2,678/11,828 | 1.00 (Ref.) | - | 154/1,612 | 1.00 (Ref.) | - | 281/7,561 | 1.00 (Ref.) | - | 85/1,398 | 1.00 (Ref.) | - |
|  | T2 | 2,781/12,918 | 1.16 (1.08,1.22) | 1.14×10^-6^ | 148/1,732 | 1.47 (1.18,1.82) | 2.21×10^-5^ | 301/7,812 | 1.23 (1.05,1.43) | 1.06×10^-4^ | 92/1,421 | 1.48 (1.14,1.82) | 2.87×10^-4^ |
|  | T3 | 3,239/12,452 | 1.23 (1.12,1.33) | 8.39×10^-8^ | 176/1,805 | 1.79 (1.43,2.33) | 8.78×10^-6^ | 369/7,904 | 1.27 (1.06,1.45) | 8.23×10^-3^ | 101/1,516 | 1.65 (1.33,2.06) | 6.01×10^-5^ |
|  | *P* for trend | 8.12×10^-8^ | | | 4.81×10^-5^ | | | 4.24×10^-3^ | | | 3.87×10^-5^ | | |

Note: HGS: handgrip strength; CVD: cardiovascular disease; SD: standard deviation; T1: the first tertile; T2: the second tertile; T3: the third tertile; HR: hazard ratio; CI: confidence interval. Continuous (standardized) and categorized (tertiles) versions of HGS were analyzed in separate models. All models were adjusted for age, education, income, drinking status, smoking status, physical activity, and BMI.

### Table S4. Association of HGS burdens with the incident CVD risk after excluding participants who developed CVD during the first two years of follow-up.

| Burdens | | UKB | | | CHARLS | | | SHARE | | | KLOSA | | |
| --- | --- | --- | --- | --- | --- | --- | --- | --- | --- | --- | --- | --- | --- |
|  |  | Events  (cases/non-cases) | HR  (95%CI) | *P*-value | Events  (cases/non-cases) | HR  (95%CI) | *P*-value | Events  (cases/non-cases) | HR  (95%CI) | *P*-value | Events  (cases/non-cases) | HR  (95%CI) | *P*-value |
| Slope | Continuous | 7,425/38,636 | 0.99 (0.94,1.05) | 0.681 | 463/4,417 | 1.37 (1.20,1.54) | 4.22×10^-7^ | 742/25,041 | 1.15 (1.06,1.25) | 8.44×10^-4^ | 278/4,348 | 1.64 (1.28,1.72) | 1.20×10^-7^ |
|  | T1 | 2,214/11,379 | 1.00 (Ref.) | - | 168/1,375 | 1.00 (Ref.) | - | 267/8,188 | 1.00 (Ref.) | - | 110/1,458 | 1.00 (Ref.) | - |
|  | T2 | 2,365/13,844 | 1.05 (1.02,1.05) | 3.21×10^-3^ | 149/1,497 | 1.35 (1.06,1.72) | 1.66×10^-2^ | 235/8,180 | 1.23 (1.10,1.47) | 3.08×10^-2^ | 98/1,430 | 1.27 (0.96,1.69) | 0.104 |
|  | T3 | 2,846/13,413 | 1.11 (1.05,1.19) | 1.403×10^-3^ | 146/1,545 | 1.75 (1.35,2.27) | 3.73×10^-5^ | 240/8,673 | 1.33 (1.11,1.61) | 3.05×10^-4^ | 70/1,460 | 2.13 (1.49,2.94) | 1.97×10^-5^ |
|  | *P* for trend | 4.17×10^-4^ | | | 8.66×10^-3^ | | | 7.71×10^-3^ | | | 2.11×10^-5^ | | |
| Cumulative | Continuous | 7,425/38,636 | 1.22 (1.11,1.32) | 1.56×10^-5^ | 463/4,417 | 1.82 (1.45,2.27) | 6.37×10^-7^ | 742/25,041 | 1.56 (1.20,2.00) | 7.24×10^-4^ | 278/4,348 | 2.44 (1.75,3.45) | 2.81×10^-7^ |
|  | T1 | 2,869/12,340 | 1.00 (Ref.) | - | 227/1,416 | 1.00 (Ref.) | - | 327/8,231 | 1.00 (Ref.) | - | 111/1,437 | 1.00 (Ref.) | - |
|  | T2 | 2,383/12,470 | 1.15 (1.01,1.30) | 4.78×10^-2^ | 140/1,454 | 1.39 (1.05,1.82) | 2.33×10^-2^ | 213/8,202 | 1.39 (1.10,1.79) | 6.04×10^-3^ | 91/1,453 | 1.56 (1.09,2.27) | 1.87×10^-2^ |
|  | T3 | 2,173/13,826 | 1.72 (1.39,2.13) | 5.65×10^-7^ | 96/1,547 | 1.59 (1.04,2.38) | 3.53×10^-2^ | 202/8,608 | 1.49 (1.03,2.13) | 3.61×10^-2^ | 76/1,458 | 2.70 (1.52,4.76) | 7/36×10^-4^ |
|  | *P* for trend | 2.23×10^-8^ | | | 5.02×10^-3^ | | | 8.44×10^-4^ | | | 7.52×10^-4^ | | |
| Relative cumulative | Continuous | 7,425/38,636 | 2.70 (2.50,2.94) | 2.58×10^-8^ | 463/4,417 | 1.37 (1.20,1.54) | 4.22×10^-7^ | 742/25,041 | 1.16 (1.08,1.27) | 8.24×10^-4^ | 278/4,348 | 1.72 (1.32,1.92) | 1.35×10^-7^ |
|  | T1 | 2,173/11,724 | 1.00 (Ref.) | - | 156/1,374 | 1.00 (Ref.) | - | 245/8,049 | 1.00 (Ref.) | - | 108/1,421 | 1.00 (Ref.) | - |
|  | T2 | 2,383/12,826 | 1.30 (1.18,1.39) | 4.68×10^-2^ | 152/1,509 | 1.39 (1.11,1.79) | 8.60×10^-2^ | 227/8,397 | 1.25 (1.14,1.52) | 2.85×10^-4^ | 92/1,460 | 1.30 (1.04,1.85) | 4.37×10^-2^ |
|  | T3 | 2,869/14,086 | 1.72 (1.39,2.13) | 6.43×10^-7^ | 155/1,534 | 1.82 (1.39,2.33) | 1.66×10^-5^ | 270/8,595 | 1.37 (1.18,1.67) | 3.01×10^-5^ | 68/1,467 | 2.33 (1.61,3.23) | 1,02×10^-5^ |
|  | *P* for trend | 3.58×10^-8^ | | | 3.43×10^-3^ | | | 5.71×10^-5^ | | | 3.16×10^-5^ | | |

Note: HGS: handgrip strength; CVD: cardiovascular disease; SD: standard deviation; T1: the first tertile; T2: the second tertile; T3: the third tertile; HR: hazard ratio; CI: confidence interval. Continuous (standardized) and categorized (tertiles) versions of HGS were analyzed in separate models. All models were adjusted for age, education, income, drinking status, smoking status, physical activity, and BMI.

### Table S5. Association of HGS transitions with incident CVD risk after excluding participants who developed CVD during the first two years of follow-up.

| Transitions | Events  (cases/non-cases) | HR (95%CI) | *P*-value |
| --- | --- | --- | --- |
| Low → low HGS | 2,816/15,762 | 1.00 (Ref.) | - |
| Low → increase HGS | 1,061/12,276 | 0.71 (0.62,0.82) | 6.23×10^-3^ |
| Moderate → moderate HGS | 872/11,823 | 1.00 (Ref.) | - |
| Moderate → high HGS | 1,254/15,624 | 0.79 (0.71,0.87) | 8.27×10^-3^ |
| Moderate → low HGS | 1,187/7,161 | 1.09 (1.03,1.15) | 7.44×10^-4^ |
| High → high HGS | 957/5,364 | 1.00 (Ref.) | - |
| High → decrease HGS | 761/4,432 | 1.11 (1.03,1.21) | 7.49×10^-3^ |

Note: HGS: handgrip strength; CVD: cardiovascular disease. HR: hazard ratio; CI: confidence interval. Continuous (standardized) and categorized (tertiles) versions of HGS were analyzed in separate models. All models were adjusted for age, education, income, drinking status, smoking status, physical activity, and BMI.

### Table S6. Association of HGS burdens with the incident CVD risk after excluding participants who underwent drug treatments at baseline and the first follow-up.

| Burdens | | UKB | | | CHARLS | | | SHARE | | | KLOSA | | |
| --- | --- | --- | --- | --- | --- | --- | --- | --- | --- | --- | --- | --- | --- |
|  |  | Events  (cases/non-cases) | HR  (95%CI) | *P*-value | Events  (cases/non-cases) | HR  (95%CI) | *P*-value | Events  (cases/non-cases) | HR  (95%CI) | *P*-value | Events  (cases/non-cases) | HR  (95%CI) | *P*-value |
| Slope | Continuous | 8,698/37,198 | 1.05 (1.00,1.10) | 0.068 | 478/5,149 | 1.41 (1.25,1.56) | 1.94×10^-8^ | 951/23,277 | 1.11 (1.04,1.20) | 3.19×10^-3^ | 278/4,335 | 1.41 (1.25,1.56) | 1.94×10^-8^ |
|  | T1 | 2,589/11,538 | 1.00 (Ref.) | - | 178/1,679 | 1.00 (Ref.) | - | 327/7,674 | 1.00 (Ref.) | - | 110/1,418 | 1.00 (Ref.) | - |
|  | T2 | 3,053/12,063 | 1.05 (0.93,1.20) | 0.474 | 159/1,626 | 1.30 (1.03,1.64) | 2.96×10^-2^ | 298/7,760 | 1.23 (1.04,1.45) | 1.52×10^-2^ | 99/1,444 | 1.27 (0.96,1.69) | 0.104 |
|  | T3 | 3,056/13,597 | 1.10 (0.96,1.27) | 0.184 | 141/1,844 | 1.85 (1.43,2.44) | 4.88×10^-6^ | 326/7,843 | 1.23 (1.05,1.45) | 1.43×10^-2^ | 69/1,473 | 2.13 (1.49,2.94) | 1.98×10^-5^ |
|  | *P* for trend | 0.462 | | | 2.47×10^-3^ | | | 3.19×10^-3^ | | | 2.14×10^-5^ | | |
| Cumulative | Continuous | 8,698/37,198 | 1.06 (1.00,1.14) | 0.080 | 478/5,149 | 1.92 (1.52,2.38) | 7.92×10^-8^ | 951/23,277 | 1.41 (1.12,1.75) | 3.25×10^-3^ | 278/4,335 | 2.44 (1.75,3.45) | 1.20×10^-7^ |
|  | T1 | 2,594/10,809 | 1.00 (Ref.) | - | 230/1,527 | 1.00 (Ref.) | - | 417/7,643 | 1.00 (Ref.) | - | 111/1,432 | 1.00 (Ref.) | - |
|  | T2 | 2,782/12,058 | 1.01 (0.91,1.12) | 0.914 | 144/1,741 | 1.43 (1.09,1.89) | 1.17×10^-2^ | 288/7,805 | 1.27 (1.11,1.33) | 1.49×10^-3^ | 91/1,437 | 1.56 (1.09,2.27) | 1.82×10^-2^ |
|  | T3 | 3,322/14,331 | 1.15 (1.00,1.33) | 0.057 | 104/1,881 | 1.56 (1.04,2.38) | 3.28×10^-2^ | 246/7,829 | 1.28 (1.14,1.37) | 5.56×10^-3^ | 76/1,466 | 2.70 (1.52,4.76) | 7.22×10^-4^ |
|  | *P* for trend | 0.062 | | | 3.01×10^-3^ | | | 3.19×10^-3^ | | | 7.37×10^-4^ | | |
| Relative cumulative | Continuous | 8,698/37,198 | 1.96 (1.85,2.08) | 1.24×10^-13^ | 478/5,149 | 1.43 (1.28,1.64) | 1.02×10^-8^ | 951/23,277 | 1.47 (1.23,1.92) | 3.25×10^-3^ | 278/4,335 | 1.49 (1.28,1.72) | 1.20×10^-7^ |
|  | T1 | 2,599/11,810 | 1.00 (Ref.) | - | 186/1,634 | 1.00 (Ref.) | - | 361/7,684 | 1.00 (Ref.) | - | 105/1,424 | 1.00 (Ref.) | - |
|  | T2 | 2,781/12,053 | 1.00 (0.90,1.11) | 0.960 | 161/1,692 | 1.33 (1.05,1.67) | 2.72×10^-2^ | 285/7,803 | 1.39 (1.04,1.45) | 2.01×10^-2^ | 102/1,442 | 1.30 (1.00,1.82) | 0.051 |
|  | T3 | 3,318/13,335 | 1.16 (1.01,1.33) | 4.60×10^-3^ | 131/1,823 | 1.92 (1.47,2.50) | 6.65×10^-3^ | 305/7,790 | 1.56 (1.15,2.17) | 1.62×10^-2^ | 71/1,469 | 2.22 (1.82,3.13) | 1.01×10^-5^ |
|  | *P* for trend | 1.01×10^-3^ | | | 1.98×10^-3^ | | | 2.11×10^-3^ | | | 1.25×10^-5^ | | |

Note: HGS: handgrip strength; CVD: cardiovascular disease; SD: standard deviation; T1: the first tertile; T2: the second tertile; T3: the third tertile; HR: hazard ratio; CI: confidence interval. Continuous (standardized) and categorized (tertiles) versions of HGS were analyzed in separate models. All models were adjusted for age, education, income, drinking status, smoking status, physical activity, and BMI.

### Table S7. Association of HGS transitions with incident CVD risk after excluding participants who underwent drug treatments at baseline and the first follow-up.

| Transitions | Events  (cases/non-cases) | SHR (95%CI) | *P*-value |
| --- | --- | --- | --- |
| Low → low HGS | 2,526/15,452 | 1.00 (Ref.) | - |
| Low → increase HGS | 1,278/13,521 | 0.74 (0.61,0.90) | 2.54×10^-2^ |
| Moderate → moderate HGS | 3,162/14,857 | 1.00 (Ref.) | - |
| Moderate → high HGS | 791/5,620 | 0.77 (0.73,0.82) | 9.62×10^-3^ |
| Moderate → low HGS | 523/8,254 | 1.10 (1.05,1.16) | 1.16×10^-4^ |
| High → high HGS | 1,234/6,364 | 1.00 (Ref.) | - |
| High → decrease HGS | 891/5,891 | 1.14 (1.06,1.23) | 3.45×10^-3^ |

Note: HGS: handgrip strength; CVD: cardiovascular disease. HR: hazard ratio; CI: confidence interval. Continuous (standardized) and categorized (tertiles) versions of HGS were analyzed in separate models. All models were adjusted for age, education, income, drinking status, smoking status, physical activity, and BMI.

### Table S8. Association of HGS burdens with incident CVD risk using Fine-Gray competing risk mortality.

| Burdens | | UKB | | | SHARE | | | KLOSA | | |
| --- | --- | --- | --- | --- | --- | --- | --- | --- | --- | --- |
|  |  | Events  (cases/non-cases) | SHR  (95%CI) | *P*-value | Events  (cases/non-cases) | SHR  (95%CI) | *P*-value | Events  (cases/non-cases) | SHR  (95%CI) | *P*-value |
| Slope | Per SD increment | 6,831/40,127 | 2.48 (1.94,3.17) | 3.69×10^-13^ | 1101/25,066 | 1.05 (0.98,1.14) | 0.180 | 277/4,049 | 1.11 (1.01,1.36) | 1.49×10^-2^ |
|  | T1 | 2,738/12,605 | 1.00 (Ref.) | - | 386/8,088 | 1.00 (Ref.) | - | 110/1,329 | 1.00 (Ref.) | - |
|  | T2 | 2,214/13,667 | 1.13 (1.06,1.20) | 1.12×10^-4^ | 345/8,282 | 1.01 (0.87,1.15) | 0.351 | 98/1,328 | 1.11 (0.85,1.44) | 0.420 |
|  | T3 | 1,879/13,855 | 1.26 (1.17,1.36) | 2.41×10^-9^ | 370/8,690 | 1.07 (0.92,1.23) | 0.199 | 69/1,392 | 1.58 (1.17,2.12) | 2.02×10^-3^ |
|  | *P* for trend | 2.50×10^-9^ | | | 0.176 | | | 1.74×10^-3^ | | |
| Cumulative | Per SD increment | 6,831/40,127 | 1.10 (1.00,1.11) | 1.04×10^-10^ | 1101/25,066 | 1.37 (1.17,1.46) | 3.13×10^-6^ | 277/4,049 | 1.20 (1.06,1.35) | 5.23×10^-5^ |
|  | T1 | 2,569/12,707 | 1.00 (Ref.) | - | 499/8,065 | 1.00 (Ref.) | - | 110/1,309 | 1.00 (Ref.) | - |
|  | T2 | 2,193/13,690 | 1.14 (1.07,1.20) | 9.86×10^-9^ | 327/8,388 | 1.72 (1.49,1.99) | 6.56×10^-5^ | 96/1,352 | 1.67 (1.23,2.26) | 2.16×10^-5^ |
|  | T3 | 2,069/13,730 | 1.37 (1.29,1.47) | 3.12×10^-9^ | 275/8,613 | 2.41 (2.01,2.89) | 3.76×10^-5^ | 76/1,388 | 2.91 (2.00,4.23) | 6.92×10^-5^ |
|  | *P* for trend | 4.56×10^-10^ | | | 4.88×10^-6^ | | | 3.54×10^-5^ | | |
| Relative  cumulative | Per SD increment | 6,831/40,127 | 1.24 (1.19,1.28) | 1.96×10^-5^ | 1101/25,066 | 1.02 (0.89,1.16) | 0.194 | 277/4,049 | 1.12 (1.04,1.36) | 2.10×10^-3^ |
|  | T1 | 2,879/12,539 | 1.00 (Ref.) | - | 386/8,174 | 1.00 (Ref.) | - | 110/1,322 | 1.00 (Ref.) | - |
|  | T2 | 2,259/13,783 | 1.15 (1.09,1.22) | 6.62×10^-5^ | 345/8,376 | 1.00 (0.87,1.15) | 0.217 | 95/1,345 | 1.11 (0.86,1.45) | 7.12×10^-3^ |
|  | T3 | 1,693/13,805 | 1.45 (1.36,1.55) | 5.23×10^-5^ | 370/8,516 | 1.07 (0.93,1.23) | 0.354 | 72/1,382 | 1.58 (1.18,2.12) | 8.45×10^-3^ |
|  | *P* for trend | 3.01×10^-5^ | | | 0.294 | | | 7.45×10^-3^ | | |

Note: HGS: handgrip strength; CVD: cardiovascular disease; SD: standard deviation; T1: the first tertile; T2: the second tertile; T3: the third tertile; HR: hazard ratio; CI: confidence interval. Continuous (standardized) and categorized (tertiles) versions of HGS were analyzed in separate models. All models were adjusted for age, education, income, drinking status, smoking status, physical activity, and BMI. No death data can be available in CHARLS, which was thus excluded from this analysis.

### Table S9. Association of HGS transitions with incident CVD risk using Fine-Gray competing risk mortality.

| Transitions | Events  (cases/non-cases) | SHR (95%CI) | *P*-value |
| --- | --- | --- | --- |
| Low → low HGS | 1,526/6,870 | 1.00 (Ref.) | - |
| Low → increase HGS | 2,194/16,976 | 0.61 (0.56,0.65) | 2.56×10^-4^ |
| Moderate → moderate HGS | 1,981/15,857 | 1.00 (Ref.) | - |
| Moderate → high HGS | 679/5,585 | 0.63 (0.57,0.69) | 7.14×10^-3^ |
| Moderate → low HGS | 1,188/8,161 | 1.49 (1.38,1.61) | 3.69×10^-5^ |
| High → high HGS | 234/6,364 | 1.00 (Ref.) | - |
| High → decrease HGS | 255/5,432 | 1.31 (1.09,1.57) | 4.12×10^-3^ |

Note: HGS: handgrip strength; CVD: cardiovascular disease. HR: hazard ratio; CI: confidence interval. Continuous (standardized) and categorized (tertiles) versions of HGS were analyzed in separate models. All models were adjusted for age, education, income, drinking status, smoking status, physical activity, and BMI.

### Table S10. Association of HGS burdens with incident CVD risk among males.

| Burdens | | UKB | | | CHARLS | | | SHARE | | | KLOSA | | |
| --- | --- | --- | --- | --- | --- | --- | --- | --- | --- | --- | --- | --- | --- |
|  |  | Events  (cases/non-cases) | HR  (95%CI) | *P*-value | Events  (cases/non-cases) | HR  (95%CI) | *P*-value | Events  (cases/non-cases) | HR  (95%CI) | *P*-value | Events  (cases/non-cases) | HR  (95%CI) | *P*-value |
| Slope | Continuous | 4,897/15,753 | 1.04 (1.00,1.08) | 0.067 | 195/2,491 | 1.37 (1.16,1.61) | 1.85×10^-4^ | 583/10,561 | 1.08 (1.00,1.16) | 0.081 | 147/1,920 | 1.39 (1.15,1.67) | 5.86×10^-4^ |
|  | T1 | 2,090/5,931 | 1.00 (Ref.) | - | 70/828 | 1.00 (Ref.) | - | 216/3,669 | 1.00 (Ref.) | - | 59/637 | 1.00 (Ref.) | - |
|  | T2 | 1,506/5,112 | 0.95 (0.77,1.18) | 0.604 | 71/823 | 1.12 (0.79,1.56) | 0.539 | 166/3,341 | 1.02 (0.88,1.18) | 0.885 | 55/630 | 1.19 (0.81,1.72) | 0.395 |
|  | T3 | 1,301/4,710 | 0.96 (0.78,1.19) | 0.682 | 54/840 | 1.75 (1.16,1.92) | 6.54×10^-4^ | 201/3,551 | 1.08 (0.93,1.25) | 0.337 | 33/653 | 2.33 (1.45,3.85) | 4.67×10^-4^ |
|  | *P* for trend | 6.21×10^-2^ | | | 7.27×10^-4^ | | | 0.336 | | | 5.66×10^-4^ | | |
| Cumulative | Continuous | 4,897/15,753 | 1.92 (1.72,1.96) | 5.02×10^-75^ | 195/2,491 | 1.85 (1.33,2.56) | 2.06×10^-4^ | 583/10,561 | 1.27 (0.96,1.64) | 0.104 | 147/1,920 | 2.08 (1.37,3.23) | 3.46×10^-4^ |
|  | T1 | 1,090/3,228 | 1.00 (Ref.) | - | 81/817 | 1.00 (Ref.) | - | 285/3,295 | 1.00 (Ref.) | - | 67/629 | 1.00 (Ref.) | - |
|  | T2 | 1,334/4,148 | 1.16 (1.02,1.32) | 3.44×10^-2^ | 68/826 | 1.16 (0.79,1.72) | 0.473 | 163/3,676 | 1.15 (0.93,1.43) | 0.206 | 47/638 | 1.30 (0.82,2.04) | 0.282 |
|  | T3 | 2,473/8,377 | 1.45 (1.23,1.45) | 7.26×10^-6^ | 46/848 | 1.64 (0.94,2.86) | 0.083 | 135/3,590 | 1.28 (0.97,1.69) | 0.087 | 33/653 | 1.72 (0.92,3.33) | 0.095 |
|  | *P* for trend | 1.11×10^-7^ | | | 0.067 | | | 8.93×10^-2^ | | | 0.095 | | |
| Relative cumulative | Continuous | 4,897/15,753 | 1.89 (1.75,2.04) | 2.10×10^-73^ | 195/2,491 | 2.33 (1.79,3.13) | 5.91×10^-4^ | 583/10,561 | 1.14 (0.99,1.20) | 0.104 | 147/1,920 | 1.47 (1.23,1.75) | 3.06×10^-5^ |
|  | T1 | 2,082/5,892 | 1.00 (Ref.) | - | 78/831 | 1.00 (Ref.) | - | 198/3,397 | 1.00 (Ref.) | - | 59/637 | 1.00 (Ref.) | - |
|  | T2 | 1,492/5,251 | 1.15 (1.01,1.32) | 4.21×10^-2^ | 77/825 | 1.19 (0.98,1.61) | 0.067 | 184/3,520 | 1.14 (0.99,1.20) | 0.075 | 55/630 | 1.19 (0.81,1.72) | 0.395 |
|  | T3 | 1,323/4,610 | 1.45 (1.23,1.69) | 7.04×10^-6^ | 51/835 | 1.47 (1.20,1.67) | 4.32×10^-4^ | 201/3,644 | 1.20 (0.95,1.27) | 0.225 | 33/653 | 2.33 (1.45,3.85) | 4.67×10^-4^ |
|  | *P* for trend | 1.02×10^-7^ | | | 0.087 | | | 8.97×10^-2^ | | | 5.66×10^-4^ | | |

Note: HGS: handgrip strength; CVD: cardiovascular disease; SD: standard deviation; T1: the first tertile; T2: the second tertile; T3: the third tertile; HR: hazard ratio; CI: confidence interval. Continuous (standardized) and categorized (tertiles) versions of HGS were analyzed in separate models. All models were adjusted for age, education, income, drinking status, smoking status, physical activity, and BMI.

### Table S11. Association of HGS burdens with incident CVD risk among female.

| Burdens | | UKB | | | CHARLS | | | SHARE | | | KLOSA | | |
| --- | --- | --- | --- | --- | --- | --- | --- | --- | --- | --- | --- | --- | --- |
|  |  | Events  (cases/non-cases) | HR  (95%CI) | *P*-value | Events  (cases/non-cases) | HR  (95%CI) | *P*-value | Events  (cases/non-cases) | HR  (95%CI) | *P*-value | Events  (cases/non-cases) | HR  (95%CI) | *P*-value |
| Slope | Continuous | 3,801/22,507 | 1.15 (1.04, 1.27) | 7.41×10^-3^ | 320/2,798 | 1.39 (1.20,1.59) | 7.88×10^-6^ | 518/14,505 | 1.07 (1.03,1.12) | 1.71×10^-3^ | 130/2,429 | 1.20 (1.08,1.35) | 3.10×10^-5^ |
|  | T1 | 1,218/6,824 | 1.00 (Ref.) | - | 113/924 | 1.00 (Ref.) | - | 170/4,414 | 1.00 (Ref.) | - | 54/781 | 1.00 (Ref.) | - |
|  | T2 | 1,334/7,413 | 1.12 (1.02,1.24) | 1.49×10^-3^ | 99/945 | 1.47 (1.11,1.96) | 7.34×10^-3^ | 179/4,955 | 1.36 (1.11,1.69) | 2.16×10^-3^ | 41/793 | 1.12 (0.94,1.33) | 0.138 |
|  | T3 | 1,249/8,270 | 1.27 (1.14,1.34) | 1.75×10^-2^ | 108/929 | 1.75 (1.30,2.38) | 3.21×10^-4^ | 169/5,136 | 1.42 (1.13,1.78) | 4.92×10^-3^ | 35/855 | 1.19 (1.01,1.41) | 7.18×10^-3^ |
|  | *P* for trend | 2.38×10^-3^ | | | 2.17×10^-6^ | | | 2.49×10^-3^ | | | 6.97×10^-3^ | | |
| Cumulative | Continuous | 3,801/22,507 | 2.70 (2.33,3.13) | 2.90×10^-11^ | 320/2,798 | 1.89 (1.43,2.50) | 3.67×10^-6^ | 518/14,505 | 2.17 (1.38,3.44) | 4.77×10^-3^ | 130/2,429 | 1.79 (1.25,2.56) | 5.39×10^-5^ |
|  | T1 | 2,012/9,433 | 1.00 (Ref.) | - | 164/873 | 1.00 (Ref.) | - | 292/4,543 | 1.00 (Ref.) | - | 67/776 | 1.00 (Ref.) | - |
|  | T2 | 1,396/8,885 | 1.20 (1.01,1.45) | 4.90×10^-2^ | 90/947 | 1.61 (1.19,2.17) | 2.10×10^-3^ | 146/4,767 | 1.06 (0.86,1.31) | 0.907 | 35/808 | 1.02 (0.83,1.25) | 6.92×10^-3^ |
|  | T3 | 393/4,189 | 1.52 (1.12,2.04) | 6.96×10^-3^ | 66/978 | 1.92 (1.25,2.94) | 2.94×10^-4^ | 80/5,195 | 1.10 (0.99,1.20) | 0.316 | 28/845 | 1.23 (0.83,1.82) | 6.82×10^-3^ |
|  | *P* for trend | 1.69×10-5 | | | 7.10×10^-6^ | | | 0.437 | | | 4.39×10^-3^ | | |
| Relative  cumulative | Continuous | 3,801/22,507 | 2.17 (1.96,2.44) | 7.50×10^-13^ | 320/2,798 | 1.43 (1.23,1.64) | 4.98×10^-6^ | 518/14,505 | 1.21 (1.08,1.36) | 2.92×10^-3^ | 130/2,429 | 1.25 (1.11,1.39) | 1.75×10^-6^ |
|  | T1 | 1115/6,674 | 1.00 (Ref.) | - | 103/931 | 1.00 (Ref.) | - | 177/4,693 | 1.00 (Ref.) | - | 51/793 | 1.00 (Ref.) | - |
|  | T2 | 1405/7,615 | 1.19 (1.00,1.41) | 6.31×10^-2^ | 101/956 | 1.52 (1.12,2.00) | 4.01×10^-3^ | 170/4,649 | 1.13 (1.08,1.36) | 3.91×10^-2^ | 44/798 | 1.19 (1.01,1.41) | 7.60×10^-3^ |
|  | T3 | 1281/8,218 | 1.59 (1.16,2.13) | 3.44×10^-3^ | 116/911 | 1.82 (1.33,2.44) | 2.39×10^-4^ | 171/5,163 | 1.23 (1.05,1.44) | 3.41×10^-3^ | 35/838 | 1.22 (1.14,1.33) | 6.57×10^-3^ |
|  | *P* for trend | 2.01×10^-5^ | | | 3.04×10^-6^ | | | 2.49×10^-3^ | | | 3.25×10^-3^ | | |

Note: HGS: handgrip strength; CVD: cardiovascular disease; SD: standard deviation; T1: the first tertile; T2: the second tertile; T3: the third tertile; HR: hazard ratio; CI: confidence interval. Continuous (standardized) and categorized (tertiles) versions of HGS were analyzed in separate models. All models were adjusted for age, education, income, drinking status, smoking status, physical activity, and BMI.

### Table S12. Association of HGS burdens with incident CVD risk among participants less than 65 years old.

| Burdens | | UKB | | | CHARLS | | | SHARE | | | KLOSA | | |
| --- | --- | --- | --- | --- | --- | --- | --- | --- | --- | --- | --- | --- | --- |
|  |  | Events  (cases/non-cases) | HR  (95%CI) | *P*-value | Events  (cases/non-cases) | HR  (95%CI) | *P*-value | Events  (cases/non-cases) | HR  (95%CI) | *P*-value | Events  (cases/non-cases) | HR  (95%CI) | *P*-value |
| Slope | Continuous | 7,100/34,593 | 1.06 (1.03,1.09) | 3.06×10^-2^ | 371/3,750 | 1.22 (1.09,1.37) | 4.08×10^-3^ | 245/11,088 | 1.05 (0.96,1.16) | 0.291 | 124/2,480 | 1.32 (1.06,1.64) | 1.36×10^-2^ |
|  | T1 | 2,118/10,111 | 1.00 (Ref.) | - | 135/1,183 | 1.00 (Ref.) | - | 65/3,980 | 1.00 (Ref.) | - | 40/708 | 1.00 (Ref.) | - |
|  | T2 | 2,328/12,880 | 0.92 (0.79,1.06) | 0.248 | 124/1,293 | 1.14 (1.00,1.23) | 0.078 | 80/3,196 | 1.10 (0.85,1.41) | 0.503 | 45/882 | 1.82 (1.25,2.70) | 2.34×10^-3^ |
|  | T3 | 2,654/11,602 | 0.93 (0.96,1.11) | 0.149 | 112/1,274 | 1.32 (1.11,1.59) | 2.56×10^-2^ | 102/3,912 | 1.11 (0.87,1.45) | 0.426 | 39/890 | 2.94 (1.85,4.55) | 3.66×10^-6^ |
|  | *P* for trend | 0.064 | | | 1.53×10^-2^ | | | 0.437 | | | 1.80×10^-6^ | | |
| Cumulative | Continuous | 7,100/34,593 | 1.64 (1.54,1.72) | 3.73×10^-53^ | 371/3,750 | 1.67 (1.27,2.17) | 6.69×10^-4^ | 245/11,088 | 1.19 (0.87,1.64) | 0.198 | 124/2,480 | 2.00 (1.25,3.23) | 4.29×10^-3^ |
|  | T1 | 2,101/9,488 | 1.00 (Ref.) | - | 181/1,137 | 1.00 (Ref.) | - | 62/3,208 | 1.00 (Ref.) | - | 50/773 | 1.00 (Ref.) | - |
|  | T2 | 2,399/12,481 | 1.14 (0.96,1.35) | 0.147 | 113/1,204 | 1.08 (1.00,1.15) | 0.176 | 87/3,698 | 0.93 (0.66,1.30) | 0.079 | 35/819 | 1.79 (1.09,2.86) | 2.18×10^-2^ |
|  | T3 | 2,600/12,624 | 1.30 (1.04,1.64) | 2.51×10^-2^ | 77/1,409 | 1.22 (1.01,1.49) | 4.66×10^-2^ | 96/4,182 | 0.93 (0.58,1.47) | 0.722 | 39/888 | 3.03 (1.43,6.67) | 4.27×10^-3^ |
|  | *P* for trend | 1.44×10^-2^ | | | 0.089 | | | 0.401 | | | 3.96×10^-3^ | | |
| Relative  cumulative | Continuous | 7,100/34,593 | 1.72 (1.54,1.92) | 5.23×10^-20^ | 371/3,750 | 1.22 (1.11,1.30) | 2.78×10^-4^ | 245/11,088 | 1.08 (0.94,1.19) | 0.367 | 124/2,480 | 1.56 (1.37,1.82) | 4.04×10^-10^ |
|  | T1 | 2,598/10,486 | 1.00 (Ref.) | - | 135/1,074 | 1.00 (Ref.) | - | 56/3,151 | 1.00 (Ref.) | - | 42/781 | 1.00 (Ref.) | - |
|  | T2 | 2,401/11,477 | 1.12 (0.95,1.33) | 0.175 | 124/1,283 | 1.05 (0.98,1.12) | 0.328 | 91/3,712 | 1.11 (0.89,1.14) | 0.620 | 50/808 | 1.54 (1.05,2.27) | 2.60×10^-2^ |
|  | T3 | 2,101/12,630 | 1.32 (1.04,1.64) | 2.43×10^-2^ | 112/1,393 | 1.20 (1.09,1.35) | 3.89×10^-2^ | 98/4,225 | 1.12 (0.91,1.16) | 0.452 | 32/891 | 2.63 (1.69,4.17) | 2.89×10^-5^ |
|  | *P* for trend | 1.48×10^-2^ | | | 5.80×10^-3^ | | | 0.225 | | | 2.23×10^-5^ | | |

Note: HGS: handgrip strength; CVD: cardiovascular disease; SD: standard deviation; T1: the first tertile; T2: the second tertile; T3: the third tertile; HR: hazard ratio; CI: confidence interval. Continuous (standardized) and categorized (tertiles) versions of HGS were analyzed in separate models. All models were adjusted for age, education, income, drinking status, smoking status, physical activity, and BMI.

### Table S13. Association of HGS burdens with incident CVD risk among participants more than 65 years old.

| Burdens | | UKB | | | CHARLS | | | SHARE | | | KLOSA | | |
| --- | --- | --- | --- | --- | --- | --- | --- | --- | --- | --- | --- | --- | --- |
|  |  | Events  (cases/non-cases) | HR  (95%CI) | *P*-value | Events  (cases/non-cases) | HR  (95%CI) | *P*-value | Events  (cases/non-cases) | HR  (95%CI) | *P*-value | Events  (cases/non-cases) | HR  (95%CI) | *P*-value |
| Slope | Continuous | 1,598/3,667 | 1.10 (0.87,1.38) | 0.407 | 199/1,484 | 1.16 (0.95,1.41) | 0.127 | 881/14,198 | 1.05 (1.00,1.11) | 0.104 | 154/1,868 | 1.69 (1.39,2.08) | 2.22×10^-7^ |
|  | T1 | 457/1,243 | 1.00 (Ref.) | - | 69/492 | 1.00 (Ref.) | - | 278/4,927 | 1.00 (Ref.) | - | 74/523 | 1.00 (Ref.) | - |
|  | T2 | 561/1,183 | 1.18 (0.91,1.23) | 0.189 | 64/497 | 1.02 (0.96,1.08) | 0.296 | 267/4,979 | 1.08 (0.94,1.22) | 0.304 | 45/667 | 1.01 (0.88,1.56) | 0.988 |
|  | T3 | 580/1,241 | 1.05 (0.97,1.14) | 0.257 | 66/495 | 1.18 (0.85,1.52) | 0.558 | 336/5,012 | 1.09 (0.96,1.25) | 0.233 | 35/678 | 1.43 (0.96,2.33) | 0.160 |
|  | *P* for trend | 0.058 | | | 0.574 | | | 0.306 | | | 0.158 | | |
| Cumulative | Continuous | 1,598/3,667 | 1.56 (1.39,1.75) | 1.31×10^-14^ | 199/1,484 | 1.41 (1.00,1.54) | 0.067 | 881/14,198 | 1.18 (0.97,1.43) | 0.201 | 154/1,868 | 3.03 (1.96,4.76) | 5.19×10^-7^ |
|  | T1 | 427/1,038 | 1.00 (Ref.) | - | 97/464 | 1.00 (Ref.) | - | 322/4,850 | 1.00 (Ref.) | - | 55/552 | 1.00 (Ref.) | - |
|  | T2 | 595/1,277 | 1.04 (0.91,1.19) | 0.602 | 54/507 | 1.10 (1.02,1.19) | 3.14×10^-2^ | 247/4,541 | 1.16 (0.99,1.37) | 0.085 | 48/656 | 1.92 (1.08,3.45) | 2.91×10^-2^ |
|  | T3 | 576/1,352 | 1.14 (0.95,1.35) | 0.169 | 48/513 | 1.16 (0.99,1.20) | 0.122 | 312/4,807 | 1.19 (0.93,1.54) | 0.174 | 50/660 | 2.17 (0.93,5.26) | 0.071 |
|  | *P* for trend | 0.103 | | | 0.078 | | | 0.104 | | | 0.068 | | |
| Relative  cumulative | Continuous | 1,598/3,667 | 1.64 (1.56,1.69) | 1.27×10^-13^ | 199/1,484 | 1.19 (1.03,1.52) | 4.23×10^-2^ | 881/14,198 | 1.08 (1.00,1.12) | 0.098 | 154/1,868 | 1.49 (1.28,1.72) | 1.27×10^-7^ |
|  | T1 | 476/1,041 | 1.00 (Ref.) | - | 69/492 | 1.00 (Ref.) | - | 278/3,927 | 1.00 (Ref.) | - | 68/541 | 1.00 (Ref.) | - |
|  | T2 | 598/1,276 | 1.03 (0.90,1.18) | 0.114 | 64/497 | 1.06 (0.99,1.12) | 0.071 | 266/4,979 | 1.08 (0.94,1.22) | 0.304 | 50/652 | 1.03 (0.67,1.59) | 0.921 |
|  | T3 | 524/1,350 | 1.15 (0.96,1.37) | 0.133 | 66/495 | 1.20 (1.04,1.47) | 4.01×10^-2^ | 337/5,292 | 1.09 (0.96,1.23) | 0.233 | 35/675 | 1.92 (1.14,3.23) | 1.49×10^-2^ |
|  | *P* for trend | 0.214 | | | 0.063 | | | 0.318 | | | 0.231 | | |

Note: HGS: handgrip strength; CVD: cardiovascular disease; SD: standard deviation; T1: the first tertile; T2: the second tertile; T3: the third tertile; HR: hazard ratio; CI: confidence interval. Continuous (standardized) and categorized (tertiles) versions of HGS were analyzed in separate models. All models were adjusted for age, education, income, drinking status, smoking status, physical activity, and BMI.

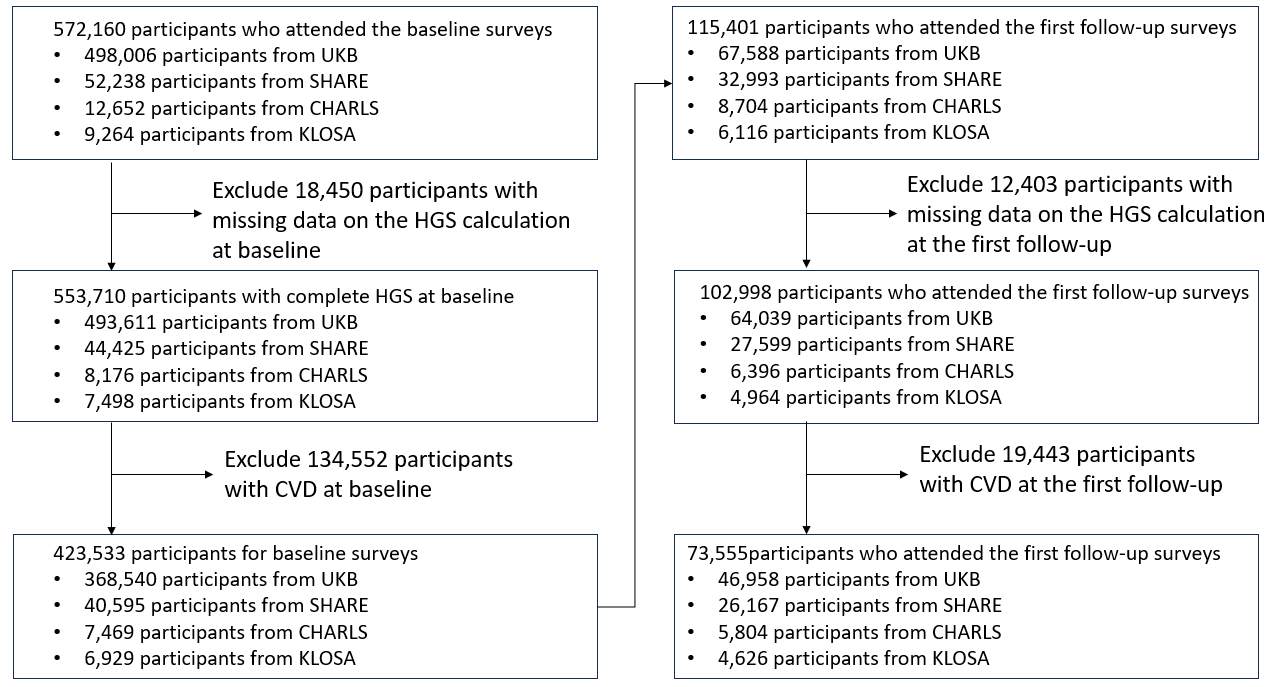

Figure S1. Flowchart of the process of sample selection in the four analyzed cohorts. HGS: handgrip strength. CVD: cardiovascular disease. Note that, new participants were recruited in the first follow-up surveys of SHARE, CHARL and KLOSA, and some previous participants also withdrew from these cohorts.

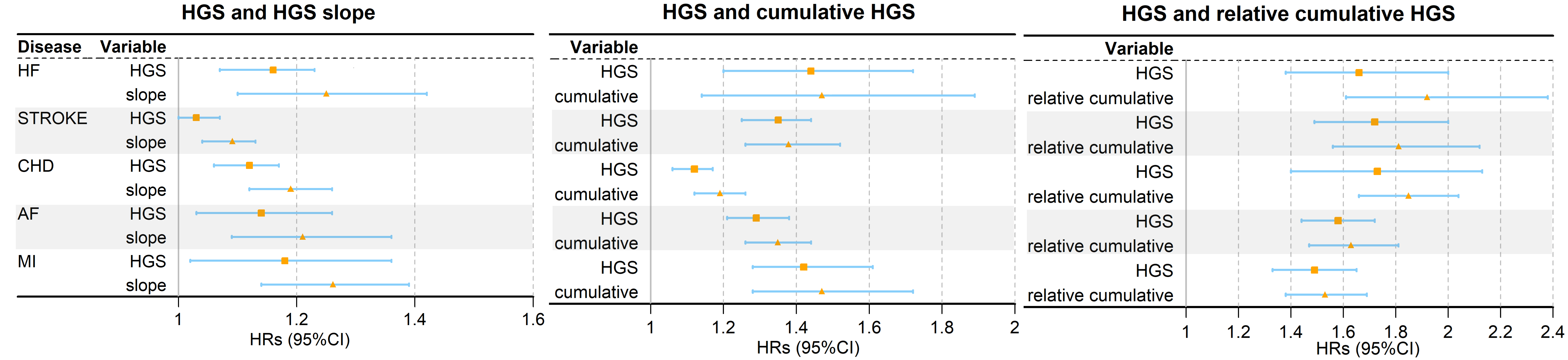
Figure S2. Forest plot illustrates the relationship between HGS and HGS burdens with individual cardiovascular disease risk in UKB. HGS: handgrip strength. HF: heart failure (295 incident cases in UKB); stroke (451 incident cases in UKB); CHD: coronary heart disease (967 incident cases in UKB); AF: atrial fibrillation (878 incident cases in UKB); MI: myocardial infarction (405 incident cases in UKB). The SHARE, CHARLS, and KLOSA cohorts did not provide the details of cardiovascular diseases as only heart and stoke problems could be available from these cohorts. As a result, this analysis was only performed in UKB.

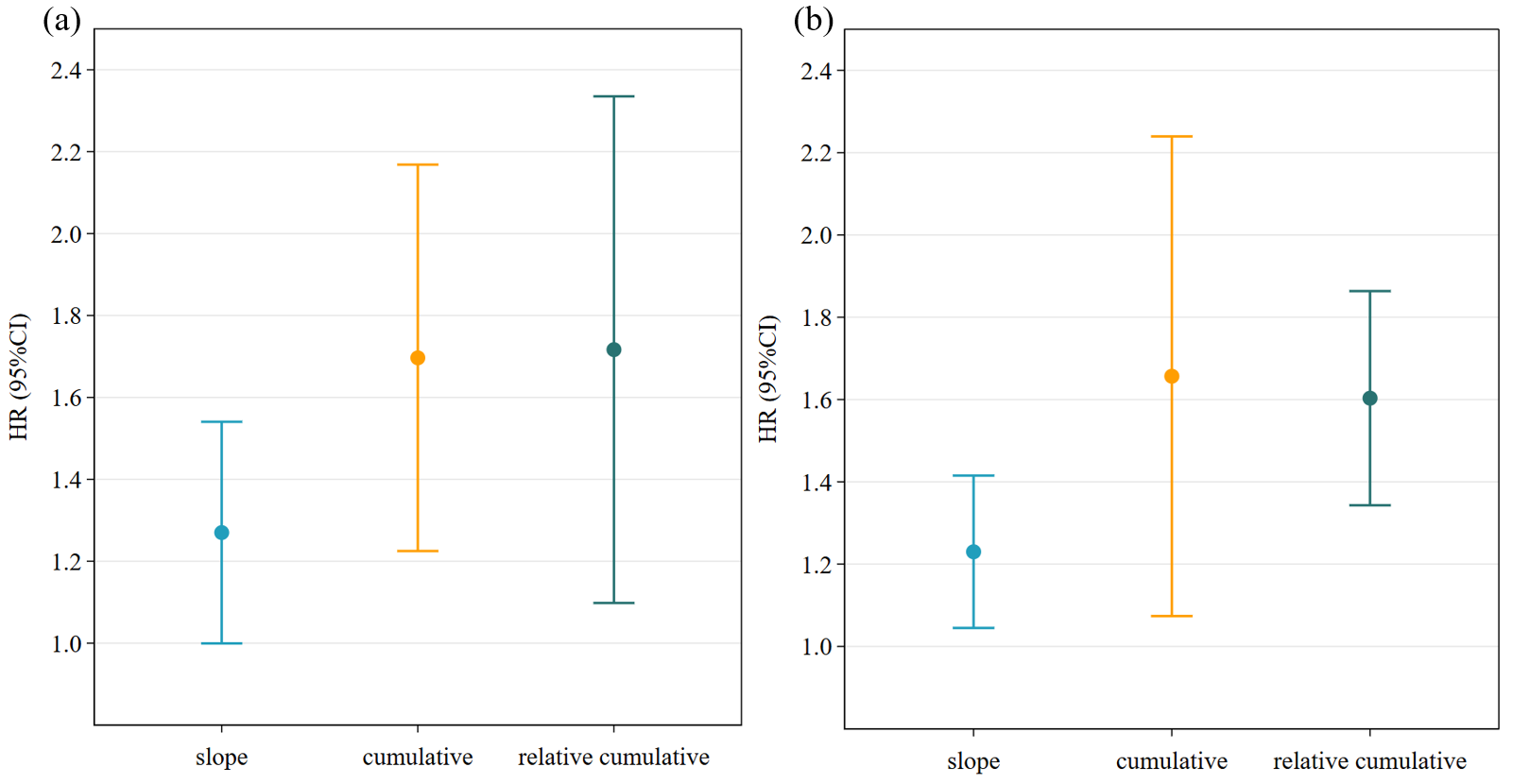

Figure S3. (a) The pooled effect of HGS burdens on the incident CVD risk after excluding participants who developed CVD during the first two years of follow-up. (b) The pooled effect of HGS burdens with the incident CVD risk after excluding participants who underwent drug treatments at baseline and the first follow-up. HGS: handgrip strength.

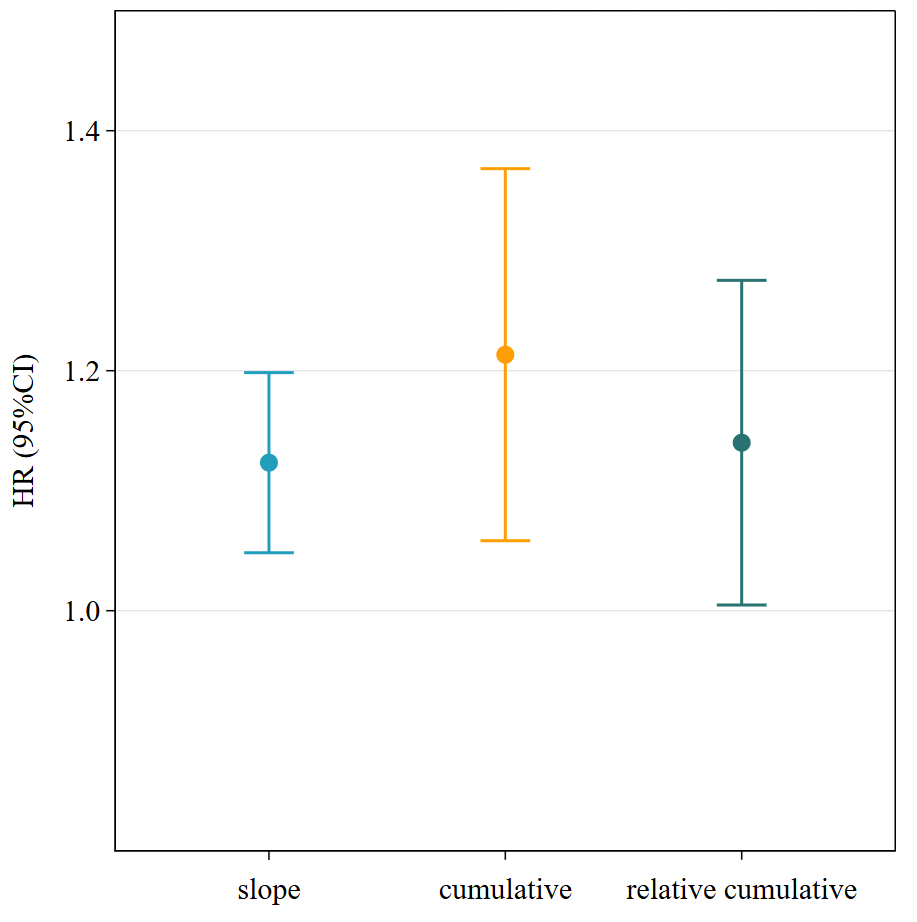

Figure S4. Association of HGS burdens with incident CVD risk after accounting for competing risk mortality. HGS: handgrip strength.

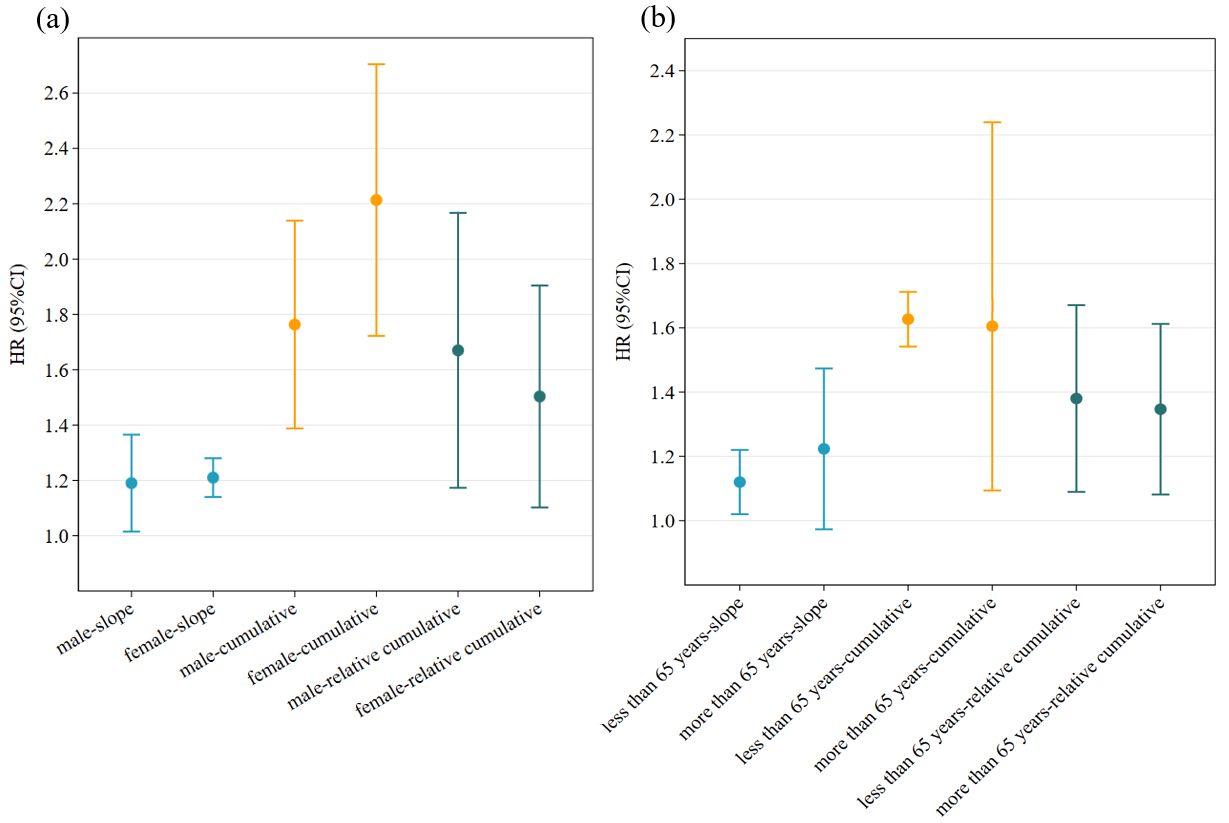

Figure S5. (a) The pooled effect of HGS burdens with the incident CVD risk stratified by sex. (b) The pooled effect of HGS burdens with incident CVD risk stratified by age. HGS: handgrip strength.

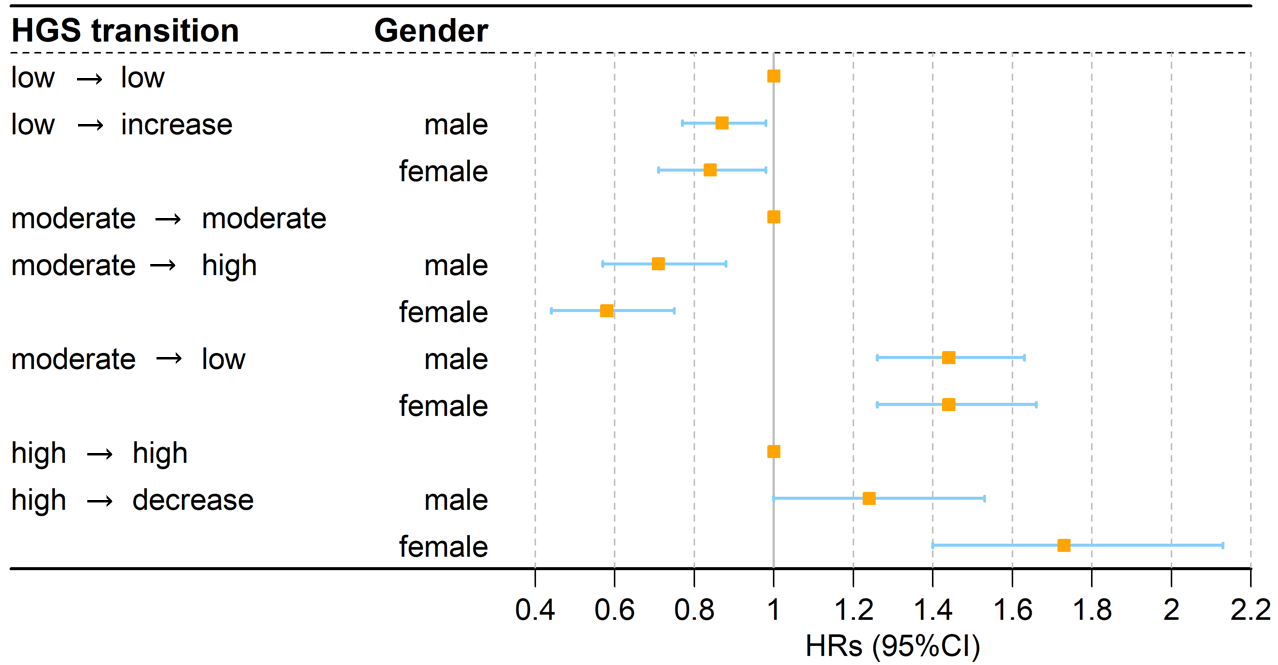

Figure S6. Forest plots of association of HGS transitions with the incident CVD stratified by sex. HGS: handgrip strength. CVD: cardiovascular disease.

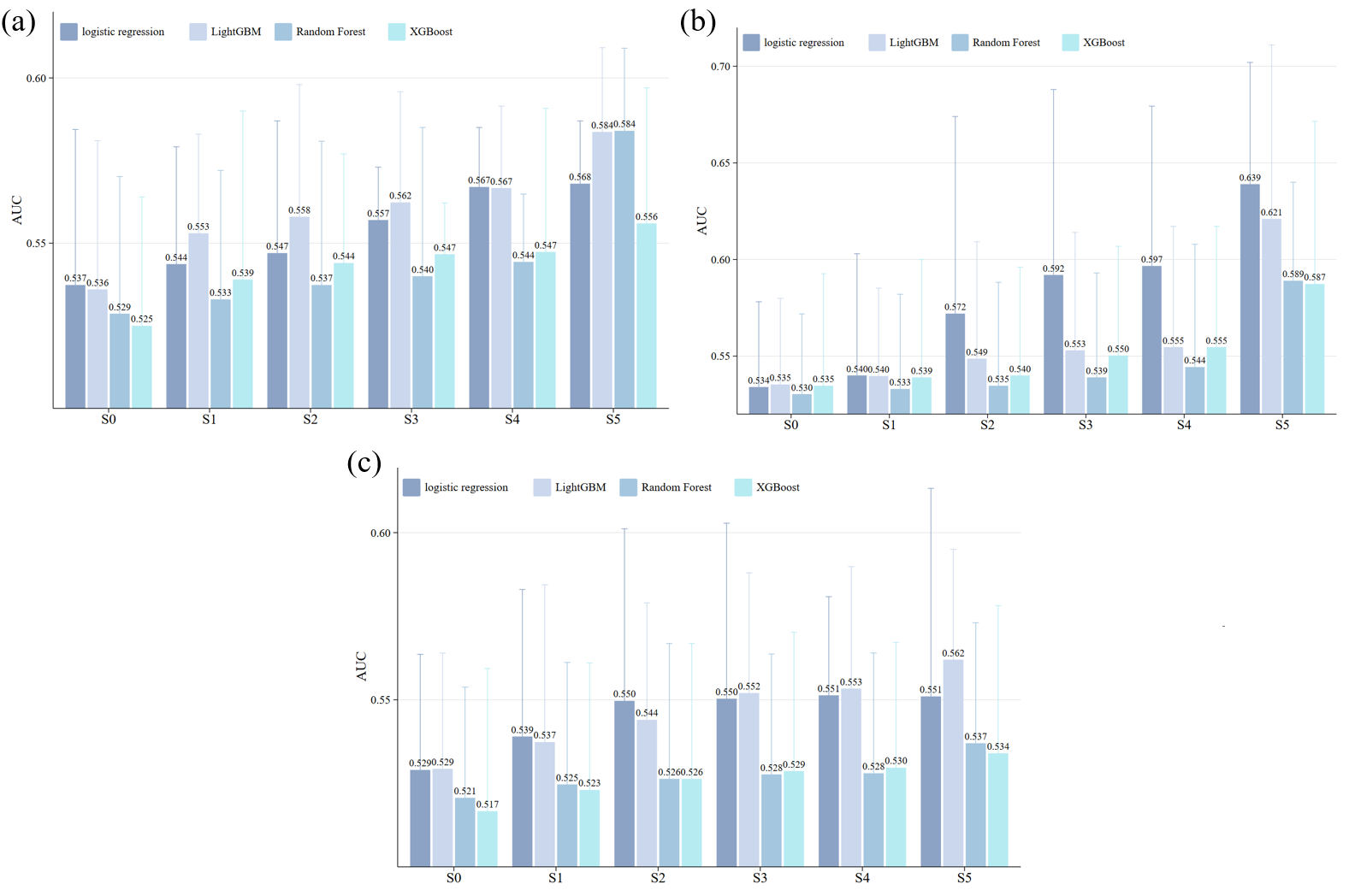

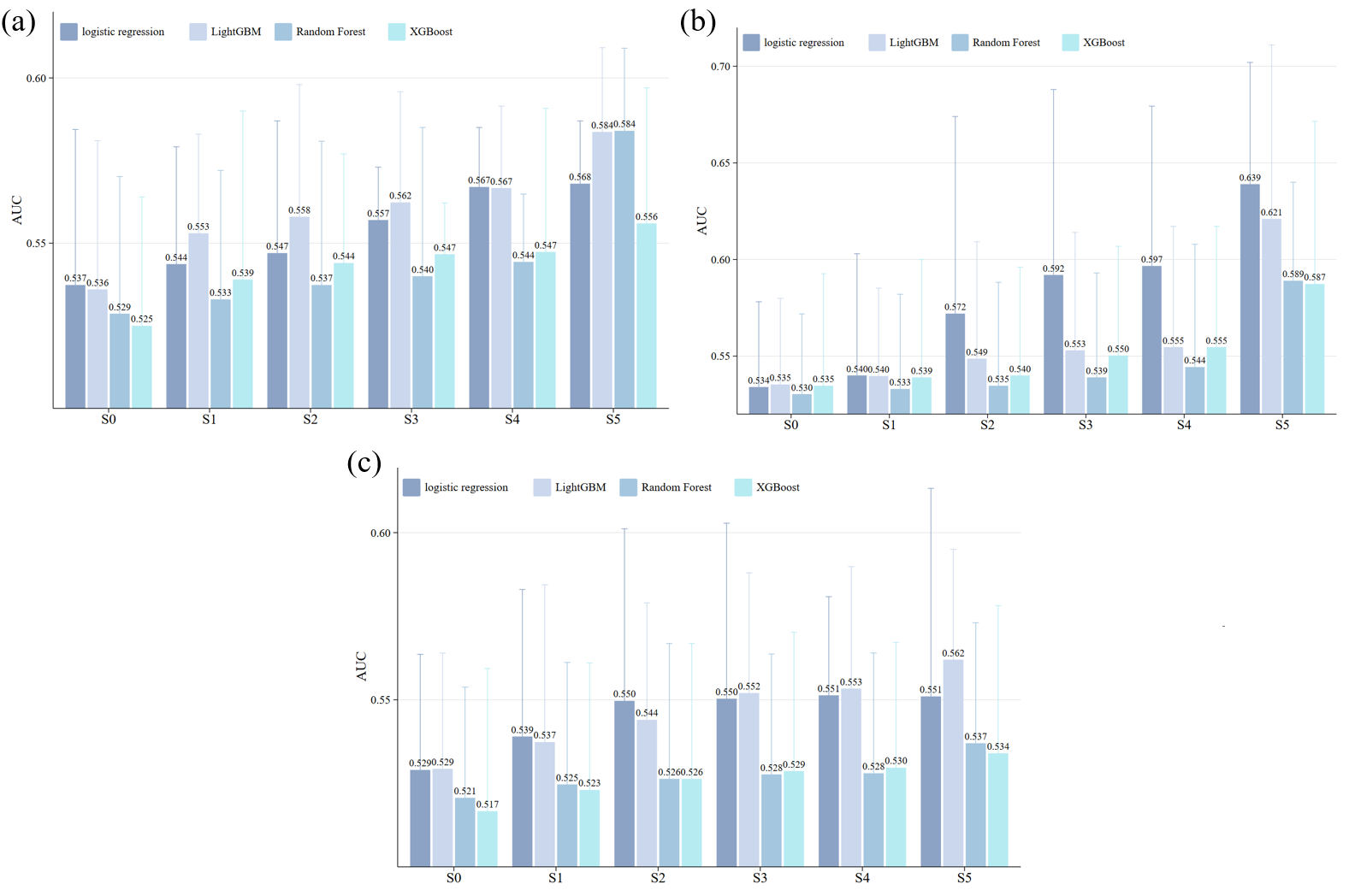

Figure S7. (a) Validated in CHARLS using models trained in European cohorts; (b) validated in UKB using models trained in CHARLS; (c) Validated in SHARE using models trained in CHARLS. Six prediction models with different sets of predictors were constructed: S0: SCORE2; S1: SCORE2 + HGS; S2: SCORE2 + HGS slope; S3: SCORE2 + cumulative HGS; S4: SCORE2 + relative cumulative HGS; S5: SCORE2 + HGS + HGS slope + cumulative HGS + relative cumulative HGS. In each model we also included covariates mentioned before, and carried out four machine learning algorithms (i.e., logistic regression, Random Forest, XGBoost and LightGBM), with the total samples randomly divided into training set (80%) and test set (20%). HGS: handgrip strength.
